# Acoustic and linguistic features of reading reveal early change, progression and function in ataxias

**DOI:** 10.64898/2026.07.10.26357775

**Authors:** Ryan Anthony J. de Belen, Yi Zheng, Matthew B. Walsh, Franziska Hoche, Chih-Chun Lin, Christopher D. Stephen, Jeremy D. Schmahmann, Lawrence White, Hasnaa Ouadoudi Belabzioui, Divya Kulkarni, Siddharth Patel, Anoopum S. Gupta

## Abstract

A major obstacle for clinical trials is the lack of objective, sensitive, and reliable measures that can detect modest changes in disease progression. Here, we determine whether acoustic and linguistic digital speech measures automatically obtained during a functionally relevant passage-reading task capture multiple dimensions of disease in ataxia, including functional communication impairment, subclinical cerebellar dysfunction and disease progression.

A total of 157 individuals with ataxia and 84 controls contributed cross-sectional data, and 54 individuals with ataxia and 43 controls contributed longitudinal data within the ongoing Neurobooth natural history study. Participants completed standardized speech recordings, patient-reported outcome measures (PROMs) and neurologist-rated clinical evaluations. A novel speech processing pipeline was developed to automatically transcribe audio recordings, identify word boundaries and extract a predefined set of linguistic and within-word acoustic features.

Individuals with ataxia exhibited marked disruption of speech timing, coordination and articulatory control, including slowed speech (*d*=1.23), prolonged inter-word pauses (*d*=-0.91), higher/more variable vocal intensity (|*d*|=0.43-0.51) and altered spectral content (|*d*|=0.43-0.79) compared to healthy controls. Linguistic features (e.g. speaking rate and within-word pause duration) showed strong associations with clinician-rated severity and PROMs (|*r*|=0.23-68), indicating alignment with functional communication impairment and patient-perceived disease burden. In contrast, acoustic features derived from cepstral measures captured subtle abnormalities in speech motor control, differentiating not only individuals with ataxia (*d*=0.65) but also pre-ataxic individuals (*d*=0.56), and those without clinically evident dysarthria (*d*=0.45), from controls. These findings indicate that acoustic features reflect subclinical cerebellar motor dysfunction involving impaired temporal coordination and vocal control before overt clinical speech impairment emerges. Longitudinally, several acoustic measures were sensitive to disease progression (*MSDR*=0.19-0.68), even in cases where clinical scales showed no detectable change. Speech-derived changes correlated with changes in clinical scales and PROMs. Both acoustic and linguistic features exhibited strong intra-session reliability.

During passage reading, acoustic and linguistic measures provide complementary but different clinical information in ataxias. Linguistic measures primarily reflect downstream functional consequences of ataxic dysarthria, whereas acoustic measures provide sensitive indicators of subclinical cerebellar motor dysfunction and progression. These findings demonstrate that natural speech analysis can produce digital measures for detecting subclinical disease, quantifying functional impairment, monitoring progression in ataxia, with strong potential for application in clinical trials and remote monitoring.

## Introduction

Cerebellar ataxias, a group of heterogeneous diseases that manifest with impaired movement and coordination,^1^ frequently present with distinctive speech abnormalities known as ataxic dysarthria.^2^ This condition was formally identified based on perceptual ratings of speech recordings obtained from individuals with cerebellar degeneration.^3^ The hallmark features of ataxic dysarthria include alterations in pitch and prosody, imprecise articulation of consonants and vowels, variability in volume, and disrupted speech rhythm and timing.^4–6^ These impairments are often accompanied by respiratory dysfunction and articulatory control deficits.^6^ Beyond their diagnostic relevance, speech disturbances substantially diminish quality of life by hindering effective communication and limiting social participation in daily activities.^7–10^ Speech abnormalities are commonly rated using clinical scales, including the Brief Ataxia Rating Scale (BARS),^11^ Scale for Assessment and Rating of Ataxia (SARA)^12^ and International Cooperative Rating Scale (ICARS)^13^. While considered gold standards, these tools are inherently subjective and prone to interrater variability.^14^ Moreover, these scales are limited by floor and ceiling effects, as well as coarse scoring granularity.^15,16^ Importantly, they were not originally designed to detect subtle or preclinical speech symptoms or monitor disease progression with high sensitivity.^14^

Disease-modifying therapies for neurodegenerative disorders, including ataxias, are rapidly advancing,^17–21^ however, a major obstacle for efficient clinical trials is the lack of objective, sensitive and reliable measures that can detect modest changes in disease progression in trials that are limited by relatively small sample sizes and short durations. Speech, however, offers a promising avenue for objective assessment.^22^ Quantitative acoustic and linguistic analyses have the potential to capture changes with finer granularity than clinician-rated scales, offering a non-invasive, scalable and repeatable measure of cerebellar function.^23^ In response, a growing body of research has focused on developing speech measures to objectively characterize the distinct speech disturbances associated with ataxic disorders,^22,24,25^ including spinocerebellar ataxias,^26–29^ Friedreich ataxia,^30–33^ ataxia telangiectasia^34^ and other ataxias^35–38^. Prior work analyzing linguistic measures during passage reading showed reduced speaking rates in ataxias.^26,28,35,39^ However, comprehensive cross-sectional and longitudinal studies systematically evaluating the potential clinical utility of speech measures during passage reading tasks in a large ataxia cohort remain limited. Passage reading tasks, which engage multiple cognitive and motor domains, provide a natural and functionally-relevant, yet standardized context for assessing speech production and enable consistent comparisons across individuals.^40^

To address this gap, we collected data in a large cohort of individuals with cerebellar ataxia diagnoses to test the hypotheses that 1) reduced speech production efficiency during passage reading is present even in the absence of clinically detectable dysarthria and 2) acoustic and linguistic measures capture multiple dimensions of disease in ataxia.

## Materials and Methods

### Data Collection

The participants and speech task analyzed in this paper were from “Neurobooth”, an ongoing natural history study collecting longitudinal multimodal data from individuals with neurodegenerative diseases.^41^ Conducted in a neurology clinic at Massachusetts General Hospital (MGH), Neurobooth involves a battery of behavioral tasks, completed in 30–40 minutes, during which synchronized multimodal data are recorded in real-time using multiple sensors. Clinician ratings and patient-reported outcome measures (PROMs) are also collected, with visits scheduled immediately before or after participants’ neurology appointments. Here, speech recorded during the passage-reading task was analyzed. Participants first viewed a brief instruction video and then read the *Bamboo Passage* (see Supplementary Material)^42–44^ aloud at their normal volume and pace, taking as much time as needed to finish the task.

### Demographic and Clinical Information

Data were collected between 28 April 2022 and 3 June 2025. Controls were individuals who were family and/or caregivers of patients who received care at the MGH Department of Neurology. Additional controls were included through a recruitment mechanism which included flyers, the Neurobooth website and advertisements posted on the Rally for Partners platform Written informed consent was obtained from the participants according to the Declaration of Helsinki. The study was approved by the MGH Institutional Review Board (2021P000257, approved March 3, 2021)

### Clinical Measures

Ataxia severity was evaluated using BARS^11^ and SARA^12^ which assess multiple domains, including gait, sitting, standing, limb coordination, speech and oculomotor function. Total scores range from 0 to 30 for BARS and 0 to 40 for SARA, with higher scores indicating greater severity. The speech subcomponents of the Modified ICARS (mICARS) were also utilized to quantify the fluency, clarity and coordination of speech.^11^ In addition to clinical assessments, a battery of PROMs was completed. The PROM-ataxia^45^ speech subscore was the mean of Items 12 and 13, motor component was the mean of Items 26 to 53, the communication subscore was the mean of Items 13, 61 and 64, and the total score was the sum of all the Items in PROM-ataxia. Dysarthria Impact Scale (DIS)^7^ and Communicative Participation Item Bank (CPIB)^46^ were also completed. Higher PROM-Ataxia scores and lower DIS and CPIB scores reflect more severely affected function.

### Automatic Speech Processing Pipeline

A fully automated speech processing pipeline was developed for transcription and feature extraction. The pipeline used a wav2vec 2.0-based model that automatically identified spoken words and their corresponding onset and offset times via forced alignment with the predicted transcript.^47^ To analyze the speech characteristics of the study participants, the following group of speech-derived features were extracted: (i) linguistic features capturing speech timing and coordination, such as speaking rate and mean pause duration, and (ii) acoustic features reflecting articulatory control, including frequency-, cepstral-, chroma-, spectral-based and higher-order measures (e.g. first- and second-order derivatives^48^). Linguistic features were derived directly from the identified words and their temporal boundaries. All acoustic features were extracted exclusively from segments contained within the detected word boundaries, such that pauses or other unvoiced intervals were excluded from the analysis. Finally, session-level summary statistics were generated by computing the mean and standard deviation for each feature, resulting in a total of 248 speech-derived features. Based on their intercorrelated nature, an aggregate summary feature was constructed separately for cepstral and chroma by taking either the average or standard deviation of all or a subset of each feature set. For example, μMFCCσ was calculated using the mean of the standard deviations of all Mel-Frequency Cepstral Coefficients (MFCCs). Further methodological details are provided in the Supplementary Methods.

### Inclusion/Exclusion Criteria

Individuals with a confirmed diagnosis of ataxia are included in the study. Healthy controls were not formally age- or sex-matched but were frequently family members of individuals with ataxia and therefore generally of similar age. No exclusion criteria were applied regarding denture use, orthodontic appliances, or other factors that may influence speech production. Information on pre-existing conditions that could affect speech, including dyslexia and other reading disabilities, were collected and documented, although these factors were not used as exclusion criteria. For correlation analysis, we excluded sessions with missing clinical assessments or with a time interval of 30 days or more between the clinical evaluation and the corresponding Neurobooth visit.

Issues relating to transcription and alignment of speech are well-documented,^49,50^ notably worsening in recordings from individuals with speech impairments.^51^ While best-performing automatic speech recognition (ASR) models like wav2vec 2.0 achieve a word error rate (WER) below 4% in typical speech,^47^ speech difficulties of our cohort required a stricter threshold to mitigate the potential influence of transcription errors on the present analyses. We excluded sessions from both groups if they fell outside a 90–116 word range, a buffer corresponding to twice the expected WER (8%) for the 98-word *Bamboo Passage*. Detailed breakdown of WER distribution, example transcripts of both the included and excluded sessions, along with participants’ corresponding disease severity and qualitative descriptions of the recorded speech are presented in Supplementary Methods.

### Acoustic-Linguistic Index for Ataxic Speech (ALIAS) Model

The Acoustic-Linguistic Index for Ataxic Speech (ALIAS) was generated using least absolute shrinkage and selection operator (LASSO) regression.^52^ Given the high dimensionality of the overall feature set, multiple subsets of acoustic and linguistic features were evaluated to predict the BARS total score, resulting in 14 trained ALIAS models. The BARS total score, rather than BARS speech score, was used because total score and speech score are strongly correlated, but BARS total has more granularity and sensitivity to reflect early-stage disease severity and may be less prone to inaccurate assessments as it is a composite score. The final ALIAS model selected for further analyses was the one demonstrating the strongest correlation with the BARS total. Additional modelling procedures, parameters, settings and model comparisons are described in the Supplementary Methods.

### Statistical Analyses

Group differences were assessed using Mann-Whitney U-tests^53^ with effect sizes measured via Cohen’s d and were interpreted as weak (0.2), moderate (0.5) or strong (0.8).^54^ Ataxic participants were categorized by severity based on the total BARS (mild if less than 8, moderate if between 8 and 16 and severe if greater than or equal to 16).^40,55^ Additionally, to assess the feature’s sensitivity to early speech changes, we compared individuals with no observable speech abnormalities (NOSA with BARS speech score of zero) to healthy controls. A separate analysis focused on participants with NOSA and with a SARA total score below 3, who were classified as pre-ataxic.^56^ Benjamini-Hochberg method was used to control for multiple comparisons.^57^ Adjusted p-values that were less than 0.05 were considered significant and subsequently reported. The reliability of the extracted features was assessed using a two-way mixed effect, absolute agreement, single measure intraclass correlation coefficient^58^ and implemented using three different sampling strategies that extract equal number of samples within each recording: *R*_l_ (temporal split): first half and second half of the recording, (2) *R*_2_ (randomized split): random half of the recording and (3) *R*_3_ (word-level shuffle): random shuffling of words (see Supplementary Methods 5). For longitudinal analysis, the progression rate of a feature was estimated by computing the slope of the fitted line to the individual’s longitudinal data. Wilcoxon signed-rank test was used to determine if the slope of the feature was significantly different from a hypothesized value of zero. The mean and standard deviation of the slope for each feature were computed, and the mean to standard deviation ratio (MSDR) was used to report effect size for sensitivity to change. Spearman’s correlation was used to assess the relationship between the extracted feature and clinical information.

## Results

### Study Participants

During enrolment, 166 individuals with ataxia and 84 healthy controls had cross-sectional data, resulting in 250 and 169 sessions, respectively. Based on the number of detected words, 24 sessions (11 sessions from 10 individuals with moderate ataxia, and 13 sessions from 10 individuals with severe ataxia) were excluded. Removing these sessions resulted in the exclusion of 9 ataxic individuals (3 with moderate ataxia and 6 with severe ataxia, approximately 4% of the full cohort) from all analyses. The final cross-sectional analysis included 157 ataxic participants (52 with NOSA, including 21 classified as pre-ataxic) and 84 controls, yielding 226 ataxia sessions (71 sessions from the NOSA group, including 30 sessions from pre-ataxic individuals) and 169 control sessions. For cross-sectional correlational analyses between speech-derived features and clinical measures, the following number of sessions were further removed due to missing clinical scores or too separated in time: BARS (speech:7, oculomotor:7 and total:7), SARA (speech:11 and total:18), MICARS (fluency/clarity:20 and alternating:21), PROM-ataxia (speech, motor and total:64 and communication:73), DIS:75 and CPIB:79. The resulting mean time intervals between clinical assessments and the corresponding Neurobooth visits were as follows: BARS and SARA:0±3 days, PROMs:6±6 days, DIS:6±6 days and CPIB:6±6 days.

The demographic and clinical information of ataxic individuals and healthy controls are detailed in Table 1. There was no age difference between individuals with ataxia (58.01±16.2 years old) and controls (56.56±17.6 years old). The NOSA group (62.8±17.6 years old) were slightly older than controls (*p*=0.04, *d*=-0.37). Finally, there was no age difference between pre-ataxic individuals (59.27±14.78) and controls. Participants with ataxia had BARS speech scores of 0.88±0.76 and BARS total scores of 10.32±6.00. Consecutive visits were 363.0±156.3 days apart for participants with ataxia and 217.5±84.0 days apart for controls. The five most frequent diagnoses were spinocerebellar ataxia type 3 (SCA-3: *n*=34), spinocerebellar ataxia type 27B (SCA-27B: *n*=14), Friedreich’s ataxia (FA, *n*=14), spinocerebellar ataxia type 6 (SCA-6: *n*=12) and cerebellar ataxia, neuropathy and vestibular areflexia syndrome (CANVAS, *n*=10).

**Table 1.**
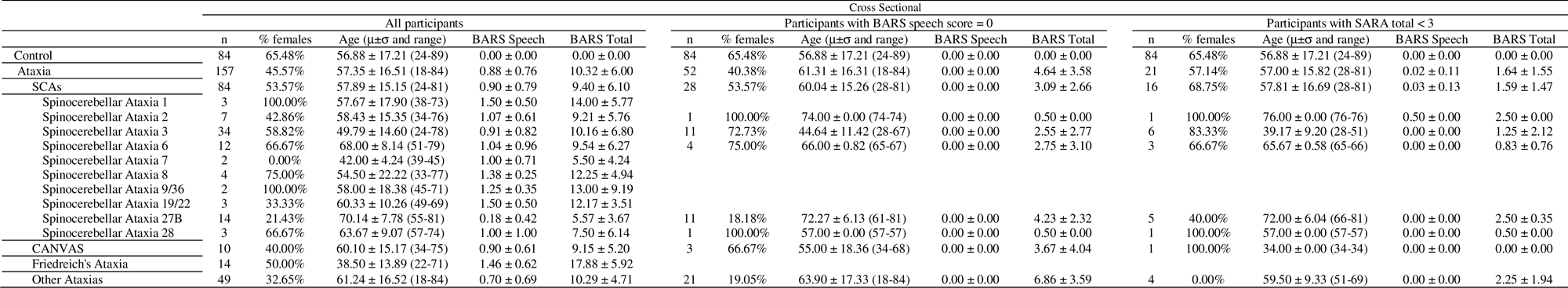
Participant Demographic and Clinical Information (Cross-Sectional Analysis)

Longitudinal data were available for 54 ataxic individuals (13 with NOSA, including 5 pre-ataxic) and 43 healthy controls, as shown in Table 2. Ataxic participants contributed an average of 1.25±0.56 years of longitudinal data, compared with 1.18±0.61 years for controls. There was no significant difference in longitudinal follow-up duration between the two groups. Restricting the analysis for the first two sessions per individual resulted in an average of 0.98±0.45 years of longitudinal data, compared with 0.61±0.27 years for controls.

**Table 2.** Participant Demographic and Clinical Information (Longitudinal Analysis)

|  | Longitudinal |  |  |  |  |
| --- | --- | --- | --- | --- | --- |
| | n | % females | Age ($\mu \pm \sigma$ and range) | BARS Speech | BARS Total |
| Control | 43 | 62.79% | 57.05 ± 18.90 (24.00-89.00) | 0.00 ± 0.00 | 0.00 ± 0.00 |
| Ataxia | 54 | 51.85% | 59.24 ± 16.34 (21.00-84.00) | 0.96 ± 0.71 | 10.94 ± 5.63 |
| SCAs | 26 | 61.54% | 58.65 ± 14.69 (24.00-79.00) | 0.98 ± 0.77 | 9.90 ± 6.31 |
| Spinocerebellar Ataxia 1 | 1 | 100.00% | 38.00 ± 0.00 (38.00-38.00) | 1.00 ± 0.00 | 7.50 ± 0.00 |
| Spinocerebellar Ataxia 2 | 4 | 50.00% | 61.50 ± 14.06 (48.00-76.00) | 0.88 ± 0.48 | 7.25 ± 4.11 |
| Spinocerebellar Ataxia 3 | 12 | 66.67% | 51.08 ± 14.43 (24.00-75.00) | 1.17 ± 0.81 | 12.79 ± 6.86 |
| Spinocerebellar Ataxia 6 | 5 | 60.00% | 70.20 ± 6.80 (64.00-79.00) | 1.00 ± 1.06 | 8.20 ± 6.64 |
| Spinocerebellar Ataxia 7 |  |  |  |  |  |
| Spinocerebellar Ataxia 8 |  |  |  |  |  |
| Spinocerebellar Ataxia 9/36 |  |  |  |  |  |
| Spinocerebellar Ataxia 19/22 | 1 | 100.00% | 63.00 ± 0.00 (63.00-63.00) | 1.00 ± 0.00 | 8.50 ± 0.00 |
| Spinocerebellar Ataxia 27B | 2 | 50.00% | 70.00 ± 0.00 (70.00-70.00) | 0.00 ± 0.00 | 3.00 ± 0.71 |
| Spinocerebellar Ataxia 28 | 1 | 0.00% | 74.00 ± 0.00 (74.00-74.00) | 1.00 ± 0.00 | 12.00 ± 0.00 |
| CANVAS | 6 | 33.33% | 66.67 ± 8.09 (52.00-75.00) | 0.83 ± 0.52 | 9.50 ± 4.10 |
| Friedreich's Ataxia | 3 | 66.67% | 43.33 ± 24.38 (25.00-71.00) | 1.17 ± 0.76 | 15.17 ± 8.95 |
| Other Ataxias | 19 | 42.11% | 60.21 ± 18.43 (21.00-84.00) | 0.95 ± 0.72 | 12.13 ± 4.22 |

**Other Ataxias:** Fragile X-Associated Tremor/Ataxia Syndrome (FXTAS), Hereditary spastic paraplegia type 7, autosomal recessive cerebellar ataxia type 1, Autosomal recessive spastic ataxia of Charlevoix-Saguenay (ARSACS), immune-mediated cerebellar ataxia, cerebellar motor syndrome, autosomal dominant cerebellar ataxia, vestibular cerebellar syndrome, Niemann-Pick Disease Type C, autosomal recessive ataxia, mild cerebellar dysfunction, cerebellar degenerative syndrome, episodic ataxia, downbeat nystagmus in primary position exaggerated by lateral gaze mild cerebellar ataxia syndrome, immune mediated ataxia syndrome, mild cerebellar atrophy downbeat nystagmus, autosomal recessive cerebellar ataxia syndrome, adult-onset slowly progressive spastic ataxia, mild cerebellar motor syndrome, Gordon-Holmes syndrome associated with mutation in PNPLA6 gene, downbeat nystagmus cerebellar ataxia, autosomal recessive cerebellar ataxia type 3, cerebellar ataxia presumed due to congenital disorders of glycosylation (CDG) syndrome, downbeat nystagmus and mild cerebellar motor findings, vestibular cerebellar syndrome of unclear etiology, cerebellar ataxia, downbeat nystagmus mild ataxia , atypical autosomal dominant episodic ataxia , Behcet's disease-associated cerebellar ataxia, pancerebellar syndrome, cerebellar atrophy right vestibulopathy, ataxia with parkinsonism and multifactorial gait disorder, prominent downbeat nystagmus appendicular ataxia, asymmetric cerebellar syndrome

### Data Quality Checks

There was no significant difference in the number of detected words between the recordings of ataxic individuals and controls. However, the word error rate (WER) differed between the two groups (*p*<0.001, *d*=-0.59, WER_ataxic_: 19.15 ± 18.89% vs WER_controls_: 8.90 ± 14.75%). Importantly, our analyses used detected word boundaries rather than the detected words themselves. Furthermore, a secondary analysis that did not exclude any sessions revealed qualitatively similar results.

### Selection of speech features

To refine the presentation of our significant results, we summarized speech-derived features based on their specific utility in either disease detection or progression tracking in Table 3. For cross-sectional analyses, features were selected for presentation if they differentiated between ataxia and controls with an effect size greater than 0.50 or distinguished both NOSA and pre-ataxic individuals from controls. For longitudinal analyses, features were selected if they demonstrated significant group-level differences in change over time or had an *MSDR*>0.40 in the ataxia group. By evaluating these criteria independently, the study avoided excluding features that may be highly informative for early detection even if they are less sensitive to progression, or vice versa. As a result, out of 248 speech-derived features, the main text focuses on a subset of 2 linguistic features and different summary statistics of 13 base acoustic features which measure articulatory control and speech dynamics. The chosen linguistic features included speaking rate and mean pause duration. The acoustic features included measures of articulatory control: MFCCs (individual components and μMFCCσ) and RMS (see Methods). Speech dynamics were quantified using spectral measures (e.g. centroid, entropy, flatness, rolloff, skewness and spread) which describe the distribution of energy across frequencies in the speech signal. These included spectral centroid, which reflects spectral brightness and spectral rolloff, which represents the frequency below which most spectral energy is concentrated. Additional measures also included chroma, crest factor, fundamental frequency (f0) contour and kurtosis. Finally, we also report the performance of the ALIAS model. Fig. 1 shows exemplars of features used in cross-sectional and longitudinal analyses grouped by participant diagnosis and severity. Comprehensive results of the performance of all speech features are provided in the Supplementary Material.

**Figure 1.**
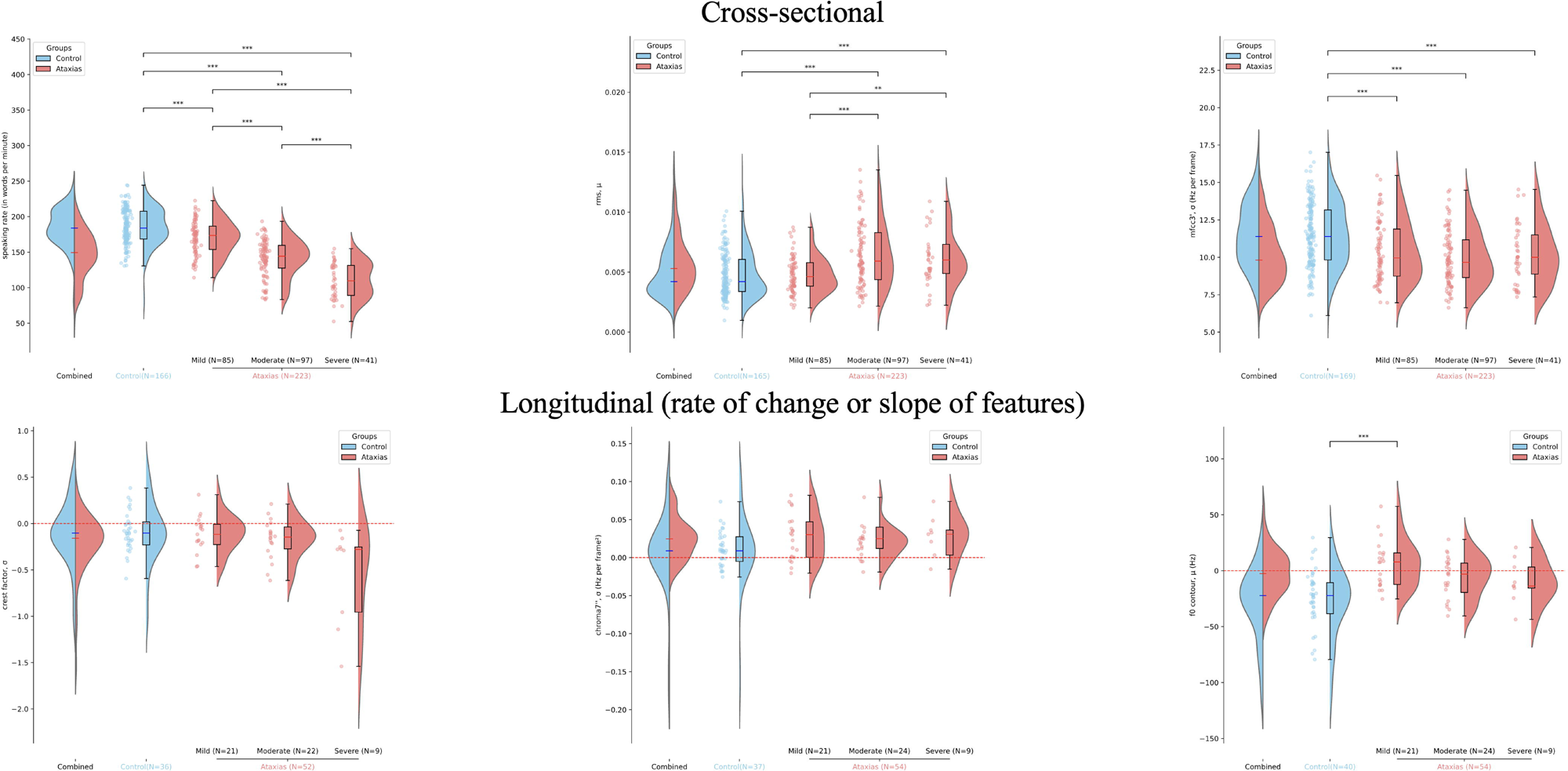
Exemplar Features used in Cross-sectional and Longitudinal Analyses grouped by participant diagnosis and severity. Each plot has (1) the comparison of the features distributions of the controls (in blue) and individuals with ataxias (in red), (2) the feature distribution of the control group, (3) the feature distribution of the individuals with ataxia with mild symptoms, (4) the feature distribution of the individuals with ataxia with moderate symptoms, (5) the feature distribution of the individuals with ataxia and severe symptoms. (2-4) show the individual scatter plot, box plot and overall shape of the feature distribution. The line bars with asterisks above each plot show the degree of significance (*P<0.05, **P<0.01 and ***P<0.001). The longitudinal plot in the second row shows the feature distribution of the slopes (rate of change of the feature) computed for each participant. Speaking rates and cepstral features (e.g. MFCC 3’, σ) are reduced in individuals with ataxia, while mean vocal intensity (RMS, μ is increased in individuals with ataxia, when compared to controls. Crest factor and chroma features are changing over time in individuals with ataxia but not controls, while f0 contour is changing over time in controls but not in the ataxia group.

**Table 3.**
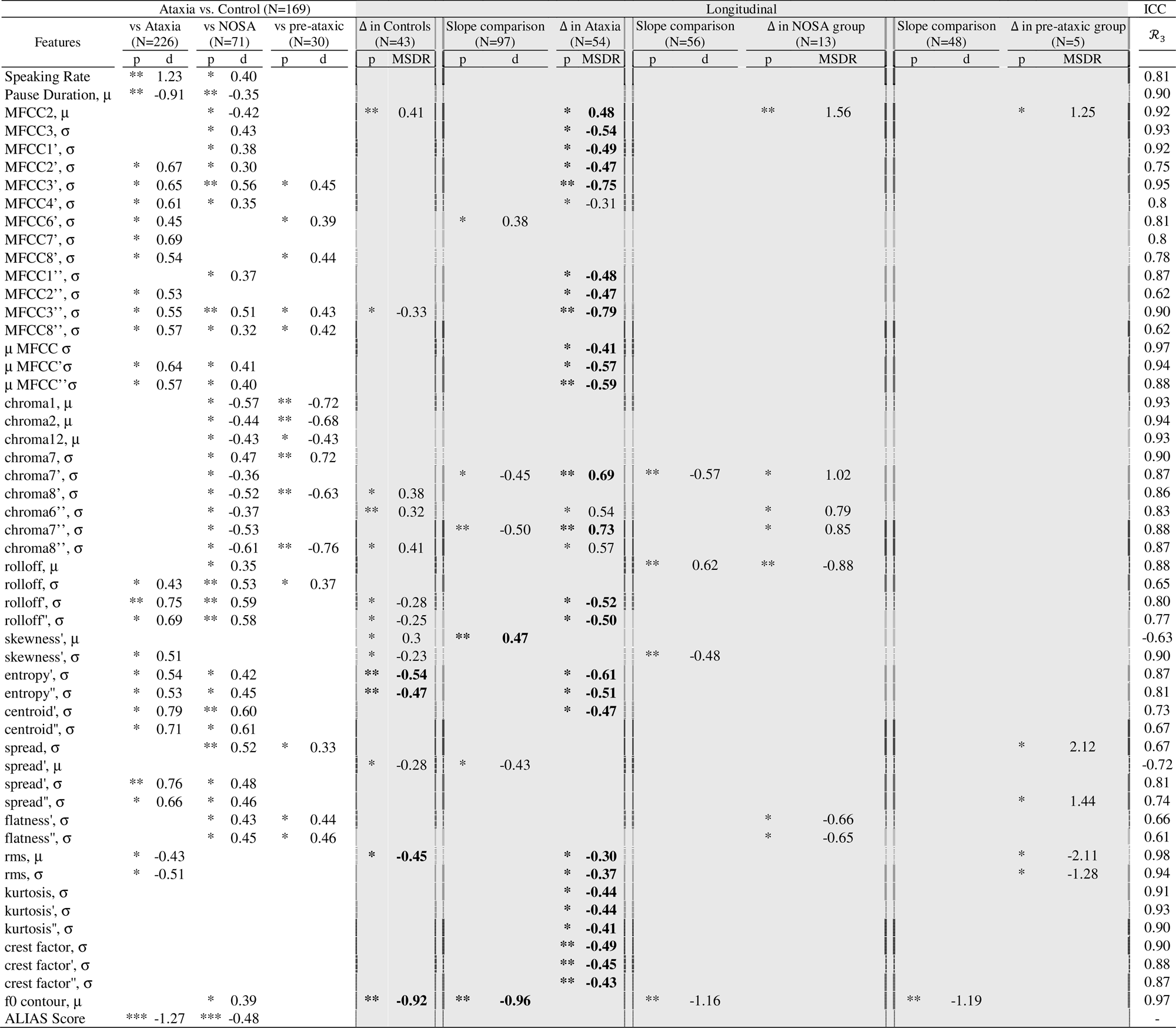
Main Results on Cross-Sectional, Longitudinal and Reliability Analysis. *All results include the p-value, Cohen’s d value, MSDR and reliability measure using interclass correlation coefficient (ICC). *P<0.05, **P<0.01, ***P<0.001. Those in bold are features that remained significant when only the first two timepoints were considered*.

### Speech abnormalities in individuals with ataxia

Individuals with ataxia exhibited reduced speaking rates and longer mean inter-word pauses compared to controls (*p*<0.01; *d*=1.23 and *d*=-0.91, respectively), as shown in Table 3. Measures of individual MFCCs (*d*=0.53-0.69) and aggregate μMFCCσ (*d*=0.57-0.64), a feature often used as a proxy for articulatory control^59^ were reduced in individuals with ataxia (*p*<0.05), reflecting impaired vocal tract dynamics. In addition, individuals with ataxia exhibited increased vocal intensity and higher irregularity in vocal intensity (RMS: *p*<0.05, |*d*|=0.43-0.51) than controls, also suggesting reduced vocal control. Finally, measures based on spectral features, such as centroid (*d*=0.71-0.79), entropy (*d*=0.53-0.54), rolloff (*d*=0.43-0.75), skewness (*d*=0.51) and spread (*d*=0.66-0.76), were reduced in individuals with ataxias (*p*<0.05), suggesting less dynamic and more monotonous speech while reading.

### Subclinical speech alterations in individuals with no observable speech abnormalities (NOSA)

Individuals with NOSA (*n*=71) exhibited slowed speech and longer mean inter-word pauses compared to controls (*p*<0.05, *d*=0.40 and *p*<0.01, *d*=-0.35, respectively), as shown in Table 3. Measures of individual MFCCs (MFCC2 μ: *d*=-0.42, MFCC σ: *d*=0.30-0.56), aggregate μMFCCσ (*d*=0.40-0.41), chroma (|*d*|=0.36-0.61) and spectral features, such as centroid (*d*=0.60-0.61), entropy (*d*=0.42-0.45), flatness (*d*=0.43-0.45), rolloff (|*d*|=0.35-0.59) and spread (*d*=0.46-0.52), were also found to be significantly different between the NOSA group and controls (*p*<0.05). The decrease in spectral features suggests that the NOSA group also experienced reduced speech dynamics and increased monotony compared to controls.

### Early speech changes in pre-ataxic individuals

Linguistic features did not differ significantly between pre-ataxic individuals (*n*=30) and healthy controls (Table 3). However, several acoustic features distinguished the two groups (*p*<0.05), including measures of individual MFCCs (*d*=0.39-0.45) and chroma (|*d*|=0.43-0.76) and spectral features, such as flatness (*d*=0.44-0.46), rolloff (*d*=0.37) and spread (*d*=0.33), indicating reduced vocal dynamics and more monotonous speech in pre-ataxic individuals.

### Reliability of speech measures during reading

As shown in Table 3, the within-session reliability of speech-derived features were consistent across different sampling strategies. For example, linguistic features exhibited high reliability (speaking rate: *R*_l_:0.81 and mean pause duration: *R*_l_:0.90. The μMFCC’σ demonstrated high reliability (*R*_l_:0.87, *R*_2_:1.00 and *R*_3_:0.94). Other speech-derived features also achieved consistent reliability (spectral entropy’ σ; *R*_l_:0.86, *R*_2_:0.98, *R*_3_:0.87, crest factor σ; *R*_l_:0.78, *R*_2_:0.95, *R*_3_:0.90, f0 contour μ; *R*_l_:0.97, *R*_2_:1.00, *R*_3_:0.97 and RMS μ: *R*_l_:0.96, *R*_2_:1.0, *R*_3_:0.98).

### Acoustic but not linguistic features capture disease progression in ataxia

Several acoustic features exhibited sensitivity to longitudinal change in the ataxia group but not in controls (Table 3). These included individual MFCCs (MFCC σ: *MSDR*= −0.31 to −0.79), μMFCCσ (*MSDR*=−0.59 to −0.41), chroma (*MSDR*=0.54−0.73), crest factor (*MSDR*= −0.49 to −0.43), spectral centroid (*MSDR*=−0.47), kurtosis (*MSDR*=−0.44 to −0.41) and RMS (*MSDR*=−0.37 to −0.34). In the NOSA group (*n*=13), several measures of individual chroma (*MSDR*=0.85-1.02) and spectral features (e.g. flatness: *MSDR*=−0.66 to −0.65 and rolloff: *MSDR*=−0.88), while in the pre-ataxic group (*n*=5), several measures of spectral spread (*MSDR*=1.44-2.12) and RMS (*MSDR*=-1.28) exhibited sensitivity to longitudinal change not observed in controls (*p*<0.05).

Linguistic features did not demonstrate significant longitudinal changes in either ataxic individuals or healthy controls Of the 29 features showing significant longitudinal change in ataxias, 22 also demonstrated significant cross-sectional differences between all ataxia, NOSA or pre-ataxic groups and controls. Among these, 20 followed trajectories consistent with clinical disease progression: 15 features that were reduced cross-sectionally in ataxic and/or NOSA participants continued to decline longitudinally, while five showed reciprocal increases, with both patterns reflecting increasing divergence from healthy baseline values over time. The remaining 2/22 features (RMS μ and σ) deviated from the expected direction of disease progression, however only showed weak cross-sectional group differences and weak longitudinal change.

When the analysis was restricted to the first two timepoints for each individual with ataxia, measures of individual MFCCs (|*MSDR*|=0.19−0.49), chroma (*MSDR*=0.37−0.47), μMFCCσ (*MSDR*=−0.31 to −0.46) and spectral rolloff (*MSDR*=−0.38 to −0.41), continued to demonstrate significant change over time (*p*<0.05). In the NOSA group, multiple individual MFCCs and chroma coefficients (MFCCs: |*MSDR*|=0.49-1.16 and chromas: *MSDR*=0.56−0.68) showed significant sensitivity to longitudinal change (*p*<0.05). This analysis restricted to the first two time points demonstrated sensitivity to disease progression over a shorter time interval.

Several acoustic features, including fundamental frequency (f0) contour (*p*<0.01, *MSDR*=−0.92) and measures of spectral features (*p*<0.05), such as spread (*MSDR*=−0.28) and skewness (|*MSDR*|=0.23−0.30), demonstrated significant changes over time in the control group whereas no changes were observed in individuals with ataxia (Table 3). When restricting the analysis to the first two timepoints, f0 contour (*p*<0.01, *MSDR*=−0.81) and measures of spectral features (*p*<0.05), such as centroid (|*MSDR*|=0.34−0.37), entropy (|*MSDR*|=0.24−0.36), kurtosis (*MSDR*=0.36) and skewness (*MSDR*=−0.28) significantly decreased in healthy controls, but not in the ataxia group. These changes may suggest habituation and increased task familiarity within the control cohort.

### Longitudinal changes in clinical measures of disease severity

The change in BARS (speech: *p*<0.01, *MSDR*=0.43 and total: *p*<0.001, *MSDR*=0.58), SARA (*p*<0.01, speech: *MSDR*=0.41 and total: *MSDR*=0.28), MICARS (clarity *p*<0.001, *MSDR*=0.41), PROM-ataxia (*p*<0.001, motor: *MSDR*=0.53 and total: *MSDR*=0.62), DIS (*p*<0.05, *MSDR*= −0.27) and CPIB (*p*<0.01, *MSDR*=−0.47) differed significantly from zero among all ataxia group (Supplementary Table 1). For the NOSA group (*n*=13), the change in PROM-ataxia (total: *MSDR*=0.69), DIS (*MSDR*=−0.56) and CPIB (*MSDR*=−0.57) were significantly different from zero (*p*<0.01). For pre-ataxic individuals (*n*=5), the change in PROM-ataxia (motor: *p*<0.01, *MSDR*=0.68 and total: *p*<0.001, *MSDR*=1.16) and DIS (*p*<0.001, *MSDR*=-1.40) were significantly different from zero. When restricting the longitudinal analysis to the first two time points, only the all ataxia group maintained significant changes from zero for all clinical scores except DIS. No significant changes were observed for the pre-ataxic or NOSA groups for the first two time points across the clinical measures (Supplementary Table 1).

### Associations between speech and clinical scores/PROMS

Linguistic features had moderate to strong associations (*p*<0.01) with multiple clinician-rated and PROMs (Table 4). Relationships with oculomotor severity were also evaluated as reading aloud involves oculomotor control. For example, speaking rate had negative associations with BARS (speech: *r*=−0.67, oculomotor: *r*=−0.23 and total: *r*=−0.67), SARA (speech: *r*=−0.63, total: *r*=−0.68), MICARS (fluency and clarity: *r*=−0.61, alternating: *r*=−0.58) and PROM-ataxia (speech and motor: *r*=−0.47, communication: *r*=−0.43, total: *r*=−0.44). It had positive associations with DIS (*r*=0.53) and CPIB (*r*=0.43).

**Table 4.**
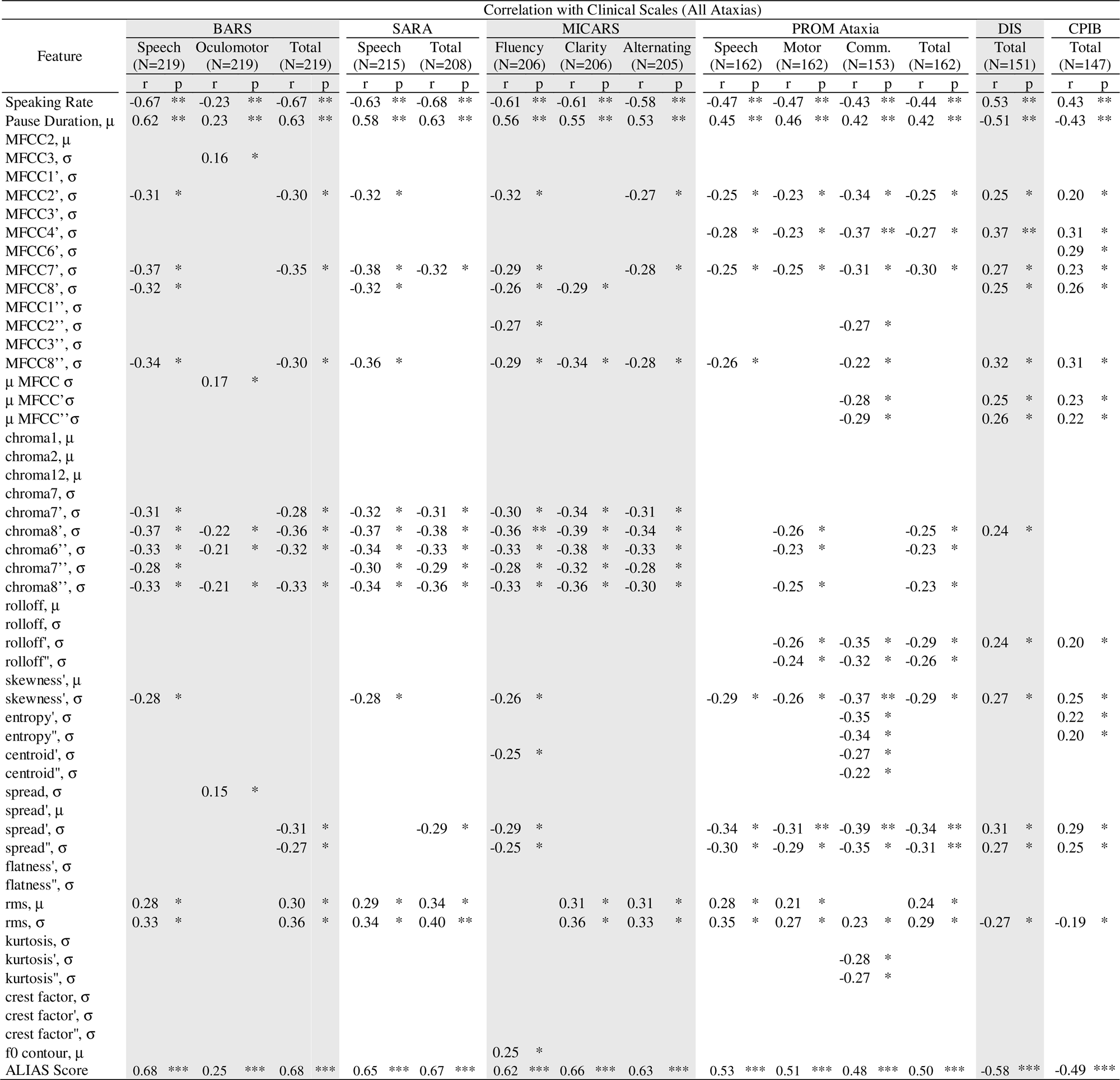
Main Results on Correlational Analysis. *All results include the correlation coefficient (r value) and P values. *P<0.05, **P<0.01, ***P<0.001*.

Acoustic features exhibited significant associations (*p*<0.05) with BARS (speech: |*r*|=0.28−0.37, oculomotor: |*r*|=0.15−0.22, total: |*r*|=0.27−0.36), SARA (speech: *r*=0.28−0.38, total: |*r*|=0.29−0.40), MICARS (fluency: |*r*|=0.25−0.36, clarity: |*r*|=0.29−0.39, alternating: |*r*|=0.28−0.34), PROM-ataxia (speech: |*r*|=0.25−0.35, motor: |*r*|=0.21−0.31, communication: |*r*|=0.22−0.39, total: |*r*|=0.23−0.34), DIS (|*r*|=0.24−0.37) and CPIB (|*r*|=0.19−0.31), as shown in Table 4.

### Associations between rate of change in speech features and clinical scores/PROMS

Rate of change of speaking rate (*r*=−0.36) and mean pause duration (*r*=0.38) were correlated with rate of change in PROM-ataxia total (*p*<0.05), as shown in Table 5. There were significant correlations between rate of change of acoustic features and rate of change of clinical measures, BARS (oculomotor: *p*<0.01, |*r*|=0.35−0.36, total: *p*<0.05, |*r*|=0.28−0.31), SARA (total: *p*<0.05, |*r*|=0.28−0.40), MICARS (*p*<0.01, fluency and clarity: |*r*|=0.29, alternating: *r*=−0.33), PROM-ataxia (*p*<0.05, speech: |*r*|=0.28−0.41, motor: |*r*|=0.28−0.39, communication: |*r*|=0.28−0.40, total: |*r*|=0.29−0.47), DIS (*p*<0.05, |*r*|=0.29−0.38) and CPIB (*p*<0.01, |*r*|=0.29−0.34). Comprehensive results are shown in Supplementary Table 10.

**Table 5.**
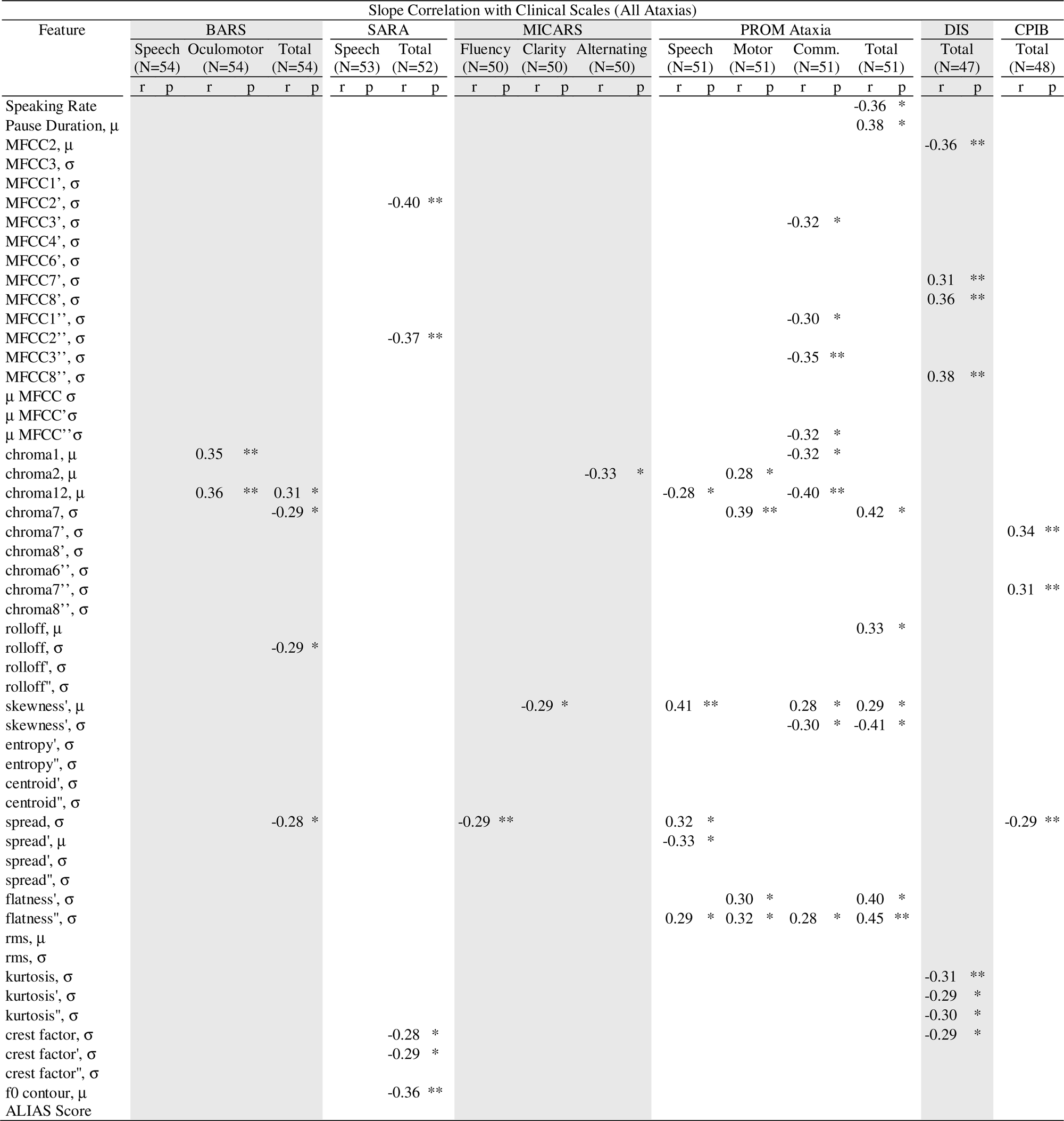
Main Results on Correlational Analysis of Slopes. *All results include the correlation coefficient (r value) and P values. *P<0.05, **P<0.01, ***P<0.001*.

### ALIAS Model Analysis

While all ALIAS models (using various acoustic and linguistic combinations) showed correlations with BARS total ranging from 0.20 to 0.68, the ALIAS model trained using all speech-derived features achieved the highest performance in BARS total estimation (*r*=0.68) and was chosen for subsequent analyses. The final ALIAS model integrates speaking rate, pause/speech durations, measures of μMFCCσ and spectral features, RMS, crest factor and f0 contour. More details on the ALIAS model training are provided in Supplementary Methods 4.

The ALIAS for ataxic participants significantly differed from controls (*p*<0.001, *d*=-1.27), a trend that persisted in the NOSA group (*p*<0.001, *d*=−0.48). However, no significant difference was observed for pre-ataxic individuals compared to controls. The final ALIAS model (and the other trained ALIAS models) was not sensitive to longitudinal changes in ataxic participants (including both NOSA and pre-ataxic individuals). ALIAS had a moderate correlation (*p*<0.001) with BARS scores (speech: *r*=0.68, oculomotor: *r*=0.25, total: *r*=0.68,), SARA scores (speech: *r*=0.65, total: *r*=0.67), MICARS (clarity: *r*=0.62, fluency: *r*=0.66, alternating: *r*=0.63), PROM-ataxia (speech: *r*=0.53, motor: *r*=0.51, communication: *r*=0.48, total: *r*=0.50,), DIS (*r*=−0.58) and CPIB (*r*=−0.49).

## Discussion

In this study, we presented a novel automated framework for characterizing and tracking speech alterations in a large and clinically diverse cohort of individuals with ataxia. Analysis of speech during a functionally relevant passage reading task revealed that linguistic and acoustic features reflect reduced speech production efficiency in individuals with ataxia, including pre-ataxic and those with no observable speech abnormalities (NOSA). We also found that acoustic and linguistic measures during reading each captured different aspects of speech dysfunction in ataxia. Linguistic measures (e.g. speaking rate and mean pause duration) which reflect speech timing and coordination are robust markers of disease severity and functional communication impairment but did not progress over time. In contrast, acoustic measures (e.g. cepstral and spectral features) which reflect articulatory control were sensitive to subclinical disease and progression. This was especially apparent during the initial two timepoints in the subclinical populations, where acoustic analysis captured subtle changes that traditional clinical assessments failed to capture. Overall, these findings support the use of digital speech measures from passage reading as objective tools for detecting early disease and sensitively tracking progression in ataxia, with potential applications in decentralized natural history studies and clinical trials.

Previous investigations have reported reduced speaking rates in individuals with SCA1,^26^ SCA2,^28^ SCAs^39^ and ARSACS^35^ relative to controls. Notably, this reduction was also observed in pre-ataxic SCA1 mutation carriers,^26^ whereas pre-ataxic individuals with SCA2 did not show this alteration.^28^ In this study, we observed reduced speaking rates in a large cohort of individuals with various forms of ataxia, indicating that slowed speech is a more general property across ataxic disorders. Among individuals with NOSA (but observable ataxia in other motor domains such as arm movement or gait), speaking rate was reduced compared with controls. Although not statistically significant, speaking rates of pre-ataxic individuals were closer to that of controls than to the ataxia group. Beyond slowed speech, ataxic individuals and those with NOSA showed prolonged inter-word pauses, longer and more irregular inter-word pauses and increased reading completion time. Together, these findings support that reduced speed of speech production is a robust and generalizable characteristic of ataxia-associated dysarthria, even at stages where abnormalities may not yet be apparent.

In terms of acoustic features, a prior cross-sectional study demonstrated that individuals with FA had slower instantaneous spectral change and a reduced spectral range during passage reading and conversational tasks compared to controls.^60^ In our broader cohort, we found that individuals with ataxia also exhibited slower instantaneous change in measures of spectral features during reading. The decrease in spectral features reflect reduced speech dynamics and increased monotony. In addition, ataxic individuals exhibited increased vocal intensity and higher irregularity in vocal intensity and signal complexity (e.g. entropy), compared to controls. These observed changes suggest increased vocal effort, phonatory irregularity, greater temporal dysregulation and presence of dysphonia associated with ‘noisiness’ or roughness in the voice. Certain acoustic features (e.g. MFCCs, chroma and spectral) were also significantly altered in both pre-ataxic and NOSA individuals relative to controls, suggesting that acoustic signatures of ataxic dysarthria are present even in the early, pre-ataxic stage of disease.

Consistent changes across linguistic and acoustic domains point to a systemic breakdown in motor speech control in individuals with ataxia. At the linguistic level, reduced speaking rate and longer pauses may reflect a compensatory “slowing down” or adaptive strategy to manage the increased demands of speech production and/or preserve accuracy and intelligibility. At the acoustic level, decreased control and accuracy manifest as increased vocal intensity, phonatory instability, imprecise articulation and altered spectral content. These findings may reflect disruption of the cerebellum’s ability to anticipate and prepare speech movements before auditory feedback arrives.^61,62^ When this predictive function is compromised, the system falls back on slower, reactive correction, resulting in the timing breakdowns and coordinative errors characteristic of cerebellar dysarthria.^2,61,63^ Together, these deficits culminate in an overall reduction in speech efficiency, defined as the ability optimize both timing and accuracy while maintaining adaptability.^64–66^ In healthy speech, this capacity depends on the cerebellum’s seamless integration of prediction, execution and real-time error correction.

While most prior work focused on clinician-rated outcomes, we also evaluated relationships between speech alterations and multiple dimensions of patient-reported disease burden. We observed robust associations between linguistic features and both clinician-rated and PROMs, extending earlier findings.^26^ As expected, speaking rate was negatively associated with ataxia severity, while longer pauses correlated with greater disease burden as captured by PROMs. Beyond these established relationships, we identified additional clinically relevant associations: greater symptom severity linked to prolonged within-word pauses, increased irregularity of inter-word pauses and longer task completion times. Linguistically, slower and more temporally dysregulated speech was associated with higher perceived impact on daily communication and quality of life, as captured by PROM-Ataxia, DIS and CPIB. Declining speaking rate and increasing pause duration were also associated with rising PROM-Ataxia scores. In contrast, acoustic features demonstrated comparatively weaker cross-sectional associations, with higher vocal intensity and increased variability in loudness showing only moderate associations with ataxia severity and overall disease burden. However, several acoustic features demonstrated significant longitudinal associations with clinician ratings and PROMs, suggesting these features reflect progressive deterioration in speech function and its functional consequences. Together, these findings highlight the relevance of natural speech production as a window into the lived experience of ataxia; as speech slows and its structure breaks down, individuals report increasing difficulties with everyday communication.

In longitudinal data analysis, we identified distinct temporal trajectories that help delineate which speech features are most sensitive for monitoring disease progression in ataxia. Prior longitudinal work reported declines in perceived speaking rate in a relatively small cohort of individuals with SCAs^67^ and reductions in spectral content in individuals with FA.^32^ In a larger ataxic cohort and using an automatic objective approach, we determined that linguistic features remained largely stable over time, potentially limiting their utility as progression markers. In contrast, acoustic features (e.g. spectral centroid, kurtosis, crest factor) demonstrated greater sensitivity to longitudinal changes. Interestingly, acoustic features exhibited similar temporal trends in both ataxic and NOSA individuals, suggesting shared speech motor profiles in early disease stages. This convergence suggests that the earliest within-word acoustic alterations may precede clinically observable behavioral manifestations, potentially identifying subclinical behavioral changes before the emergence of pronounced speech deficits. Even when limiting analysis to the first two timepoints, acoustic measures captured longitudinal changes in the ataxia population. In a small subclinical subset (NOSA=13, pre-ataxic=5), acoustic measures still captured longitudinal change despite stable clinical scales and PROMs. Sensitivity to both subclinical speech alterations and longitudinal change suggests utility as marker of disease progression with potential as quantitative endpoints in clinical trials where subtle treatment effects may not yet be reflected in conventional ratings. The use of a brief, standardized passage-reading task enables remote and repeatable assessment, making these measures well-suited for decentralized monitoring in real-world settings.

The dissociation between the properties of linguistic and acoustic features is notable. Linguistic features showed stronger cross-sectional correlations with clinical scales and PROMs but were not sensitive to change over time. Acoustic features exhibited weak cross-sectional correlations (but mildly stronger rate of change/longitudinal correlations) with clinical ratings and PROMS, but greater sensitivity to longitudinal change. Acoustic features, such as vocal intensity, spectral content and phonatory stability, are likely driven primarily by the integrity of cerebellar motor circuitry. As cerebellar dysfunction progresses, these features deteriorate in a relatively consistent trajectory within individuals, even if their absolute values vary considerably across individuals at baseline. Relatively high between-subject variability at baseline weakens cross-sectional correlations with clinical scales. However, because the within-person trajectory of acoustic change is more uniform, these features remain sensitive to longitudinal deterioration. Additionally, current clinical scales and PROMs are not designed to capture fine-grained acoustic properties, further limiting their cross-sectional correspondence while leaving longitudinal sensitivity intact. On the other hand, linguistic features related to fluency, timing and intelligibility, are more readily observable and therefore may be better captured by scales and PROMs. However, linguistic features are affected by factors beyond cerebellar dysfunction, including mood, fatigue, attention and situational variability in effort and motivation. These sources of non-disease variance may introduce noise into longitudinal measurement, masking true progression-related changes and reducing sensitivity over time. This dissociation will need to be further evaluated in future work.

Longitudinal analyses also revealed that healthy controls exhibited measurable temporal changes in acoustic features (e.g. a decrease in fundamental frequency (f0) contour, *p*<0.01, *MSDR*=−0.92), while these remained relatively stable in ataxic individuals and those with NOSA. The observed decline in f0 contour among controls may reflect motor habituation, increased task familiarity and age-related vocal changes occurring over successive sessions^68^. It is possible that their stability in the ataxia population reflect decreased familiarity, adaptation or learning over time compared with controls, consistent with the established role of the cerebellum in motor learning; however, this will need to be studied in future work.

Acoustic features, particularly MFCCs and spectral measures, showed sensitivity to both longitudinal change and subclinical disease signs. Prior research suggest that MFCC2s extracted during sustained phonation provide useful disease markers in Parkinson’s disease^59,69,70^ and Alzheimer’s disease.^71^ Evidence also suggests that MFCCs derived from running speech, particularly passage reading, capture parkinsonian deficits more effectively than those from sustained vowels.^72^ Here, we demonstrated that among the features examined, the mean of the standard deviations of all MFCCs (μMFCCσ and its derivatives) emerged as a robust marker across multiple dimensions (*p*<0.05): it distinguished ataxia from controls (*d*=0.64), identified NOSA individuals (*d*=0.41) and tracked progression (*d*=−0.57) during passage reading. Physiologically, μMFCCσ reflects the vocal tract dynamics responsible for the articulatory motions necessary for language production^59^. During reading, measures of μMFCCσ were reduced cross-sectionally in individuals with ataxia and continued to decline longitudinally. This reduction reflects decreased variability in spectral content, effectively smoothing the acoustic structure of speech and diminishing the distinct phonetic signatures of individual words. Such spectral smoothing is consistent with impaired articulatory control and increased slurring during connected speech. In contrast, during a task that does not involve articulation but requires maintaining a consistent frequency content (e.g. sustained vowel /a/ phonation, see Supplementary Results), μMFCCσ was elevated in the same cohort before declining longitudinally. Together, these findings suggest that ataxia affects speech motor control in complementary ways: reduced capacity for rapid articulatory modulation during connected speech and impaired stability in maintaining consistent vocal output during sustained phonation.

It can be beneficial to construct a single digital speech biomarker that can characterize disease phenotype, capture early signs, monitor disease progression and evaluate treatment response in clinical trials. In this study, we developed ALIAS that integrates speaking rate, pause/speech durations, measures of μMFCCσ and spectral features, RMS, crest factor and f0 contour. ALIAS differentiated ataxic individuals from healthy controls, including those with NOSA and demonstrated strong associations with clinician-rated assessments and PROMs. However, the score was not sensitive to longitudinal change, likely due to its training objective to estimate the clinical rating scale score rather than capture longitudinal changes. With ongoing longitudinal data collection, it will become possible to develop and validate a composite score trained on longitudinal data to better capture temporal patterns of disease progression.^73^

There are several limitations that should be considered when interpreting the results of this study. Reduced speaking rate exhibited by individuals with ataxia may partly reflect conscious or subconscious compensatory strategies encouraged in speech therapy rather than disease severity alone. In this study, we did not perform a subset analysis comparing those who received speech therapy to those who did not. We assumed linear relationships between speech and overall ataxia severity, as well as a linear trajectory of disease-related speech progression over time. Given the exploratory nature of this study, a broad array of linguistic and acoustic features was analyzed. Multiple comparison corrections ensured statistical robustness but narrowed the set of significant features, and confirmatory analyses on independent datasets are needed. While the overall cohort was large for a rare disease, subtype-specific sample sizes were small, limiting subgroup analyses. The pre-ataxic cohort was similarly small and findings require replication. PROM-Ataxia, DIS and CPIB contained missing data despite email and phone reminders from study staff, largely due to the need to complete surveys at home before or after the Neurobooth session given limited time on the clinic day. However, the available data (∼70%) was sufficient to evaluate relationships between speech features and patient reported function. Control participants were self-reported and did not receive formal clinical evaluation. The ASR model was trained on speech from healthy individuals, reducing accuracy for participants with more severe ataxic dysarthria, resulting in the exclusion of certain sessions due to poor word detection and a slight inclusion bias toward individuals with milder symptoms. For the severe patients that were included in the analyses, inaccurate word boundaries likely persisted, potentially affecting the quality of the extracted speech features. However, a secondary analysis that included all participants and sessions, regardless of the word detection rate, still produced consistent results, supporting the validity of our findings. Nevertheless, this limitation underscores the need for future work to refine ASR models to ensure that speech-derived features remain robust across the full spectrum of dysarthria severity.

In summary, in a large ataxia cohort, we found that digital speech measures automatically derived from a reading aloud task were able to detect early, subclinical speech changes and sensitively capture disease progression over time. The pattern of speech changes present in ataxias reflected the inherently meaningful concept of decreased efficiency. These findings demonstrate the strong potential for digital measures of naturalistic speech to be used as sensitive outcome measures and support incorporation of reading tasks across natural history studies in ataxia for further validation and development.

## Data Availability

The data are not publicly available because speech recordings contain information that could compromise the privacy of research participants.

## Acknowledgements

The authors would like to thank past members of the Neurobooth team that contributed to data collection and software development. The authors also thank the Neurobooth participants for their valuable time and feedback.

## Supplementary material

Supplementary material is available at *Brain* online

