## Supplementary Material for "Acoustic and linguistic features of reading reveal early change, progression and function in ataxias"

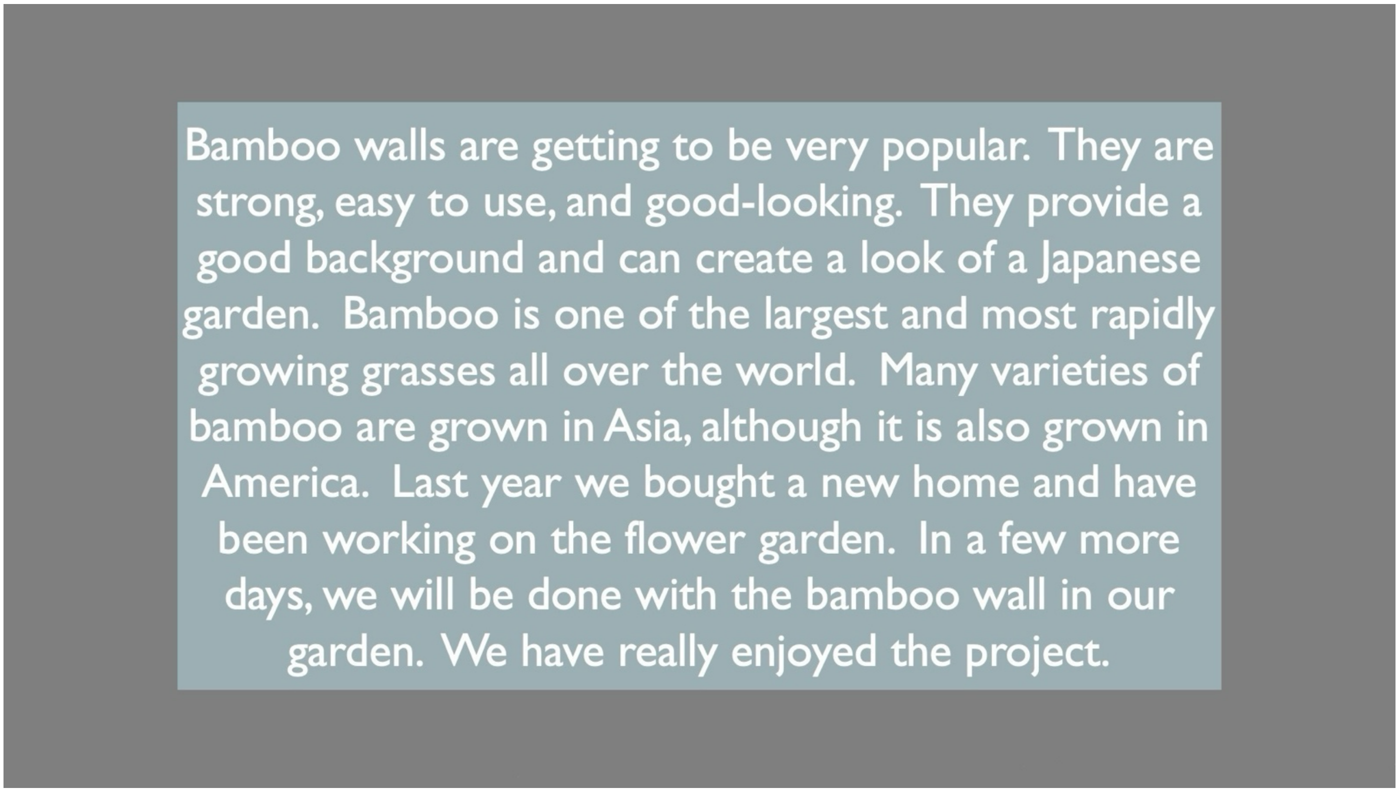

*Supplementary Material 1 The Bamboo Passage shown to the participants during the passage reading task*

#### Supplementary Tables

| Clinical Scales | | All ataxias | | | | Ataxias without OSA | | | | Pre-ataxic | | | |
|---|---|---|---|---|---|---|---|---|---|---|---|---|---|
| | | All timepoints | | First two timepoints | | All timepoints | | First two timepoints | | All timepoints | | First two timepoints | |
| | | p | MSDR | p | MSDR | p | MSDR | p | MSDR | p | MSDR | p | MSDR |
| BARS | Speech | ** | 0.43 | * | 0.38 | | | | | | | | |
| | Oculomotor | | | | | | | | | | | | |
| | Total | *** | 0.58 | ** | 0.45 | | | | | | | | |
| SARA | Speech | ** | 0.41 | * | 0.40 | | | | | | | | |
| | Total | ** | 0.28 | * | 0.22 | | | | | | | | |
| MICARS | Fluency Clarity Alternating | *** | 0.41 | * | 0.32 | | | | | | | | |
| PROM-Ataxia | Speech Motor Comm. | *** | 0.53 | ** | 0.20 | | | | | ** | 0..68 | | |
| | Total | *** | 0.62 | ** | 0.31 | * | 0.69 | | | *** | 1.16 | | |
| DIS | Total | * | -0.27 | | | ** | -0.56 | | | *** | -1.40 | | |
| CPIB | Total | * | -0.47 | * | -0.33 | | | | | | | | |

*Supplementary Table 1 Change in Clinical Measures*

### Supplementary Methods

#### Supplementary Methods 1. Speech Processing Pipeline

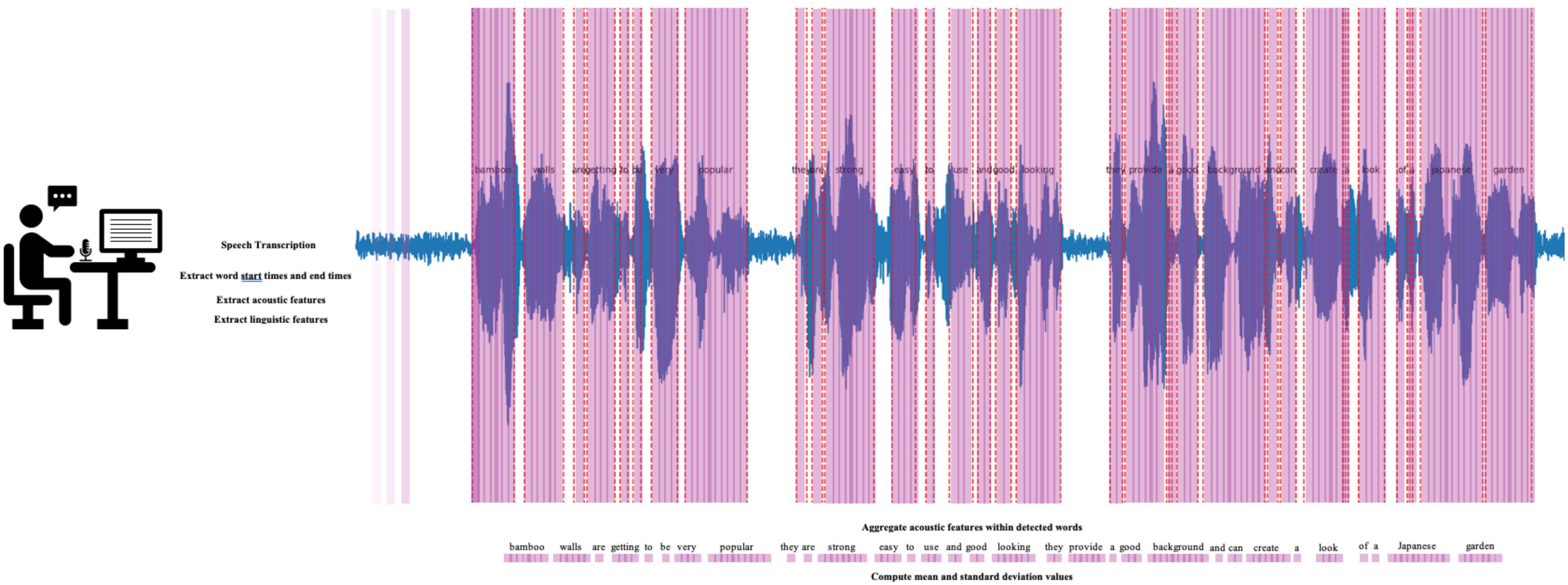

*Supplementary Figure 1 Speech Processing Pipeline*

We developed a novel, fully automated speech processing pipeline for transcription and feature extraction. The speech processing pipeline is illustrated in Supplemental Figure 1. Audio recordings, originally acquired at a sampling rate of 44.1 kHz, were resampled to 16 kHz using torchaudio (Yang et al. 2022). Afterwards, all recordings were transcribed using an automatic speech recognition (ASR) system based on the wav2vec 2.0 architecture (Baevski et al. 2020) model fine-tuned on 960 hours of English language collected from the internet. This model is among the best performing speech recognition models and achieves human-level performance. Word onset and offset times were derived using the same model via forced alignment with the predicted transcript. The detected word boundaries served as the basis for all subsequent feature extraction. Linguistic features were computed from the temporal structure defined by word onsets and offsets, with pauses operationally defined as silent intervals between consecutive words.

- Linguistic features were calculated based on the word beginning and end points. We define pauses as the stops in between words.

- Acoustic features were extracted using Surfboard (Lenain et al. 2020), an open-source Python library purposely built for clinical speech analysis. We utilized a fast Fourier transform with a 40 ms window length and 10 ms hop length to compute 13 Mel-frequency cepstral coefficients (MFCCs), 12 chroma features, and various spectral descriptors. Additional acoustic features, such as crest factor, root mean square (RMS), kurtosis, and Shannon entropy were also calculated using the same parameters, while fundamental frequency (F0) contours were estimated with a hop length of 10 ms. Based on the mentioned parameters, the pipeline automatically extracts all acoustic features across the entire session recording, generating a sequence of frames containing 'raw' acoustic features. To capture temporal dynamics, the pipeline calculated first and second derivatives from the resulting sequence of frames. The first derivative was computed as the frame-to-frame difference, quantifying the instantaneous change in 'raw' acoustic features. The second derivative was calculated by repeating this process on the results of the first derivative calculation. To ensure the analysis focused strictly on vocalized content, features were post-filtered to include only frames falling within detected word boundaries, effectively excluding pauses and unvoiced intervals. Finally, session-level summary statistics were generated by computing the mean and standard deviation of each feature across all valid speech segments within a recording.

| Group | Symbols | Features | Description | Physiological Evidence |
|---|---|---|---|---|
| Linguistic Features | | | | |
| | Speaking Rate | | The number of words divided by the total duration of the task | Reflects the pace stability and impaired timing and coordination of speech |
| | Pause Duration, μ | | The average duration between words during passage reading | Captures the difficulty in speech initiation and timing |
| | Pause Duration, σ | | The variability of duration between words during passage reading | Captures the difficulty in speech initiation and timing |
| | Percent Pause | | | |
| | Pause Events | | The number of pauses in the recording | |
| | Total Speech Duration | | The voiced segment of the speech | Captures the slowness of speech |
| Frequency Features | | | | |
| | F0 Contour | | A measure of the changes of fundamental frequency over time during speech production | Reflects the pattern of controlled changes in vocal fold vibration (*Physiological Factors Causing Tonal Characteristics of Speech: From Global to Local Prosody*, n.d.) |
| Cepstral features | | | | |
| | MFCC-n | n-th Component of Mel Frequency Cepstral Coefficients | Set of features extracted from audio signals that represent the short-term power spectrum of a sound in a way that approximates human hearing. The MFCCs assigned in the lower coefficients quantify the amplitude and spectral envelope, while those in the higher coefficients quantify information about harmonic components. | A measure of instability by capturing subtle articulatory movements of individual vocal tract elements, including the tongue and the lips (Tracey et al. 2023). |
| | μ MFCC σ | Mean of the standard deviation of all MFCCs | It is defined as the mean of the standard deviations of all MFCCs. | A summary measure of the instability of an individual's vocal tract. |
| Spectral Features | | | | |
| | slope | | A measure of the rate at which the energy of the spectrum changes with increasing frequency | Reflects the resonant characteristics of the vocal folds |
| | flux | | Rate of change (or variability) of the spectral content of a signal and reflects shifts in energy distribution over time | |
| | entropy | | A measure of the degree of randomness in the spectral distribution. It also measures the 'regularity' or 'peakiness' of the signal's power spectrum, indicating how the energy is uniformly distributed across different frequencies | Reflects the disorderliness of speech produced during passage reading |
| | centroid | | A measure that depicts the central frequency of the extracted signal, allowing it to be used as an indicator of the perceived brightness and sharpness of sound | Reflects the compromised ability to consistently articulate the words in the passage |
| | spread | | A measure of the distribution of frequencies around the mean of the spectrum | |
| | skewness | | Quantifies the asymmetry around the mean of the spectrum distribution. | |
| | kurtosis | | Defines the measure of flatness of the distribution around the mean value of the spectrum. | |
| | flatness | | a measure of uniformity of the frequency spectrum. It is the ratio of the geometric mean to the arithmetic mean of the spectrum | |
| | rolloff | | Determines the frequency below which 95% of the signal energy is contained | |
| | RMS | | The energy contained in the speech signal | Reflects the vocal intensity during passage reading |
| | crest factor | | The ratio of the peak amplitude of a signal to its RMS value. A high crest factor indicates the presence of significant peaks relative to its average value | Reflects the difficulty modulating the loudness of voice |
| Chroma Features | | | | |
| | Chroma-n | n-th Component of Chroma feature | Represents the tonal or harmonic content of the audio | |
| Higher-Order Features | | | | |
| | *feature*' | First derivative of *feature* value | A metric quantifying the difference between two temporally adjacent speech-derived features. | |
| | *feature*'' | Second derivative of *feature* value | A measure capturing the rate of change of slope between temporally adjacent speech-derived features. | |

*Supplementary Table 2 More information about the speech-derived features*

#### Supplementary Methods 2. Computing a surrogate measure for multi-dimensional features

Several acoustic features, such as mel-frequency cepstral coefficients (MFCCs) and chroma features, consist of multiple coefficients (13 coefficients for MFCC and 12 values for chroma) that capture different aspects of the acoustic signal but may also be highly intercorrelated. To evaluate redundancy within these multi-dimensional features, covariance matrices were computed to identify correlated components. As shown in Supplementary Table 3 and Supplementary Table 4, the mean values of the MFCCs and chroma features (and their higher-order derivatives) demonstrated weak correlations, whereas their standard deviation values showed moderate to strong correlations. These findings suggest that dimensionality reduction can be achieved by aggregating highly correlated features into an aggregate measure during model training. As detailed Supplementary Methods 3, two aggregation strategies were evaluated: (i) averaging all standard deviation values and (ii) incorporating only the first standard deviation values.

MFCC

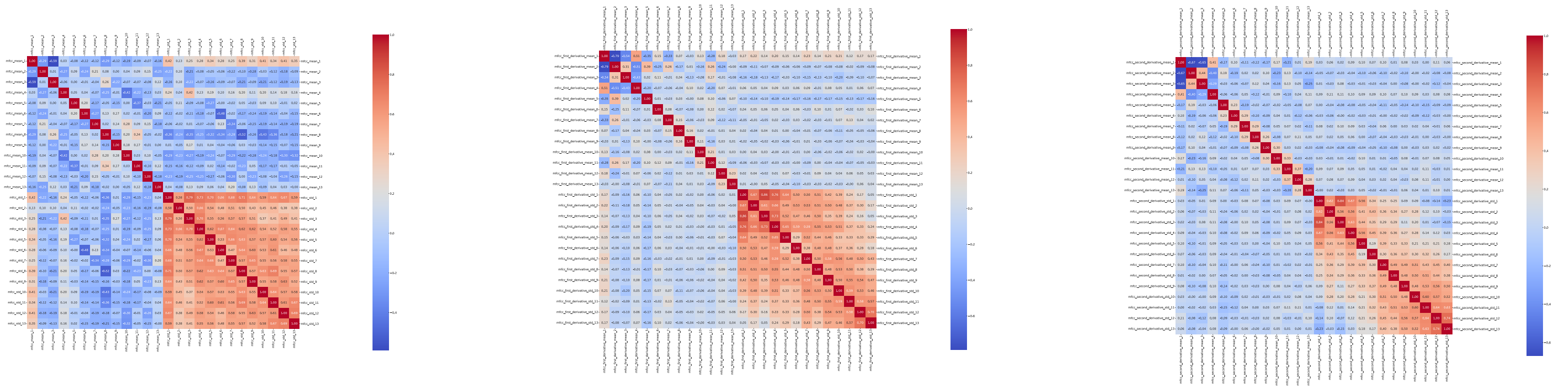

*Supplementary Table 3 Correlation Matrices of MFCCs*

Chroma

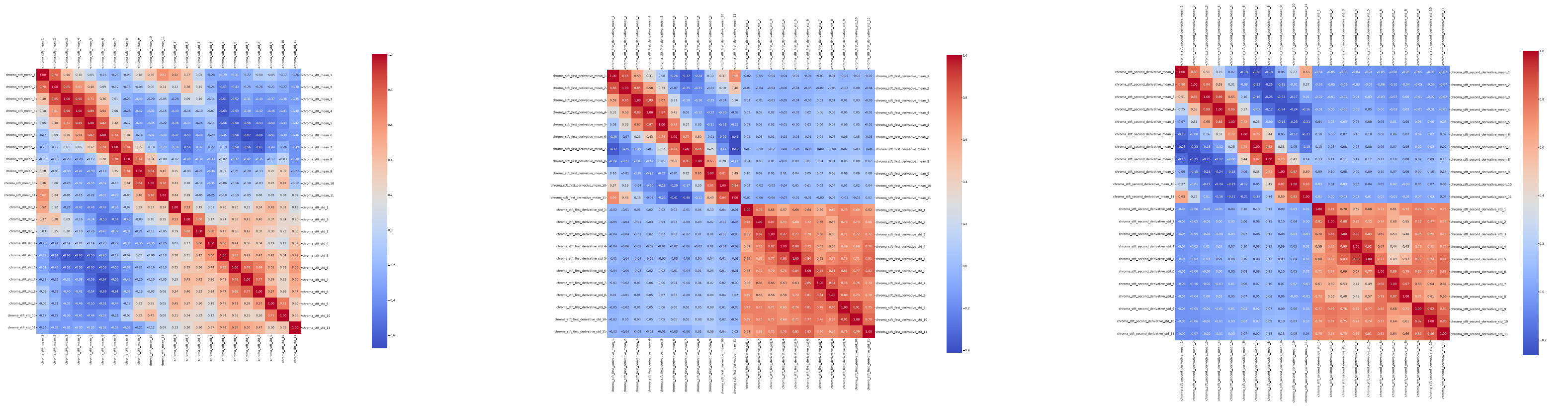

*Supplementary Table 4 Correlation Matrices of Chroma Features*

#### Supplementary Methods 3. Analysis of ASR Performance

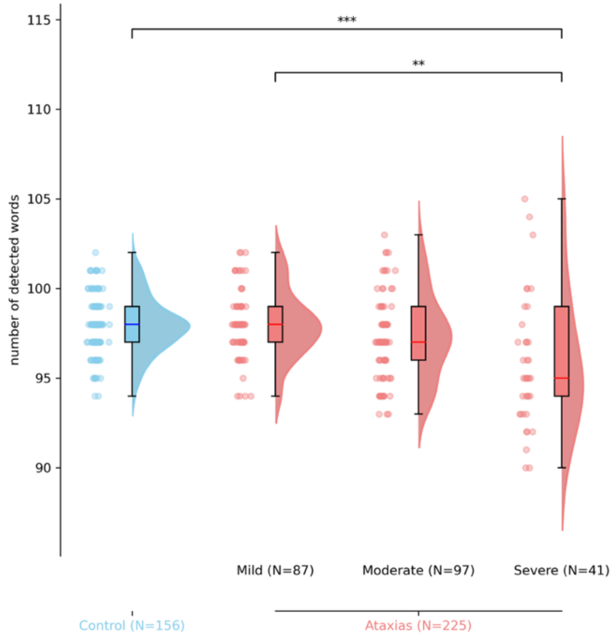

*Supplementary Figure 2 Comparison of number of detected words based on severity*

Supplementary Figure 2 highlights that the ASR performed significantly differently in participants with severe ataxia compared to healthy controls. We conducted detailed manual inspections of plots that overlaid model-generated word boundaries and detected words onto the speech waveform, as shown in Supplementary Figure 1. This review showed that the model produced accurate word boundary estimates overall, but its performance degraded in recordings with substantial unintelligibility, typically due to faint speech or pronounced slurring (22/395 sessions) and in recordings contaminated by persistent background noise when the booth was left open (2/395 sessions). Without excluding any sessions, a secondary analysis revealed qualitatively similar results. In particular, the significant effects observed in the primary analyses remained unchanged, and the associated p-values were further reduced, indicating that the results are robust even when accounting for potential inaccuracies in word detection.

| **Example of Included Transcripts** | Severity | Description of the audio file |
|---|---|---|
| bamboo walls are getting to be very popular they are strong easy to use and good looking they provide a good background and **crcan** create a look of a japanese garden bamboo is one of the largest and most rapidly growing grasses all over the world many varieties of bamboo are grown in asia although it his also grown in america last year we bought a new home and have been working on the flower garden in a few more days we will be done with the bamboo wall in our garden we have really enjoyed the project | Control | The recorded speech was intelligible. The participant hesitated and repeated the word 'can' (in **bold**). |
| bamboo walls are getting to be very popular they are strong easy to use and good looking they provide a good background and can create **a good** a look of a japanese garden bamboo is one of the largest and most rapidly growing grasses all over the world many varieties of bamboo are grown in asia although it is also grown in america last year we bought a new home and have been working on the flower garden in a few more days we will be done with the bamboo wall in our garden we have really enjoyed the project | Control | The recorded speech was intelligible. The participant misread the line and repeated the word 'a good' (in **bold**). |
| bamboo walls are getting to be very popular they are strong easy to use and good looking they provide a good background **and they** and can create a luk of a japanese garden bamboo is one of the largest and most rapidly growing grasses all over the world many varieties of bamboo are grown in asia although it is also grown in america last year we bought a new home and have been working on the flower garden in a few more days we will be don with the bamboo wall in our garden we have really enjoyed the project | Control | The recorded speech was intelligible. The participant made a mistake by adding the words 'and they' (in **bold**) during the recording. |
| bamboo walls are getting to be very popular they are strong easy to use and good looking they provide a good background and can create a woork of **(a)** japanese garden bamboo is one of the largest and most rapidly growing grasses all over the world many varieties of bamboo are grown in asia **althuogh** although it is also grown in america last year we bought a new home and have been working ond the flower garden an a few more days we will be done with the bamboo wall in our garden we have really enjoyed the project | Control | The recorded speech was intelligible. The participant missed the word **'a'** and repeated the word **'although'** |
| bamboo walls are getting to be very popular they are strong easy to use and good looking they provide a good background and can create a look of a japanese garden bamboo is one of the largest and most rapidly growing grasses all over the world many varieties of bamboo are grown in asia although it is also grown in america last year we bought a new home and have been working on the flower garden in a few more days will be don with the bamboo wall in our garden we have really enjoyed the project | Moderate | The recorded speech was intelligible, |
| bamboo walls are getting to be very popular they are strong easy to use and good looking they provide a good background and can create a look of a japanese garden bamboo is one of the largest and most rapidly growing grasses all over the world many varieties of bamboo are grown in asia although it is also grown in america last year we bought a new home and have been working on the flower garden in in a few more days we will be done with the bamboo wall in our garden we have really enjoyed the project | Moderate | The recorded speech was intelligible. |

| | | |
|---|---|---|
| bamboo walls are getting to be very popular they are strong easy to use and good looking they provide a good background and can create a look of a japanese garden bamboo is om the largest and most rapidly growing grasses all over the world mean if varieties of bamboo are grown in asia although it is also grwn in america last year we bought a new **elb e hup** but working on the flowery garden in a few more days well be done with the bamboo wal in our garden we will have really enjoyed the project | Moderate | The recorded speech was intelligible but the participant yawned while reading the seventh line of the passage. This was evident (in **bold**) in the transcription. |
| bamboo walls are getting to be very popular they are strong easy to use and good looking they provide a good background and create a look of a japanese garden bamboo is one of the largest most rapidly growing grasses all over the world many varieties of bamboo are growing in asia although it is also grown in ameriga last year we bought a new home and have been working in the flower garden in a few more days we will be ready with the bamboo pole in our garden we have really enjoyed the project | Severe | The recorded speech was intelligible. |
| bamboo walls are getn to be very popular they are strong easy and good looking they'v provide a good background and great allo of jod japanese garden bamboo is one of the largest and most rapidly growing grasses all over the world many briis a bamboo have grown in asia although it is also grown in america last year we bought a new home and have been working on the flower garden afin a few more days wewill be done with the bamboo wall in our garden and we really enjoyed the project | Severe | The recorded speech was slurred. Some words were combined or mispronounced. |
| babo walls are getting to be tat we popular they are strong easy to use and good looking they ave provided good background and can create a look of a japanese garden bambo is one of the largest and most rapidly grown grasses all over the world many varieties of bambo ar grown in asia although it is also grown in america last year we **brought** bought a new home they have been workin on the flower garden in a few more days we will be done with the bambo bal in our garden we hve really enjoyed the project | Severe | The recorded speech was slurred. The participant misread the word 'bought' and said 'brought' (in **bold**) instead. Both words were captured in the recording. |
| **ALL Excluded Transcripts** | Severity | Description of the wav files |
| bamboo walls are getting to be very popular they are strong easy to use and good looking they provide a good background and can reatea wok of the japanese garden bamboo is on of the largest and most rapidly growing grasses all over the world bany varieties of bamboo are grown in asia although it is also grown in america last year we bought a new home and have been working on the flower garden in a few more days we will be done with the bamboo wall | Mild | The participant did not finish the task completely. The recording did not include the final words *"in our garden. We have really enjoyed the project"* |
| te av e wovs are genegibe ther go ey ares troeusen us to these fees in goothey a the be fitego a gn a gan riniin the oajabenas garden a gins onov he largest amovs are gon grasses alovo bhe wv mn n enas o ao rn an an ato o ow aes amos gron an oarba ass gerboay new home on av an gring on the fourgraran a few more days me vi be done wi t baovan the god gn av ilann doithe progecten | Severe | The recorded speech was faint. The participant was struggling to perform the task. There are a few repetition of words. |
| baboo walls are getting to be fair popular they are strong easy to use and good looking tbvout a good background and create a look lik japanese garden bambou is one of the lagest most properly growing grasss all over the world many varieties of babou a gonaya although a so born in america last year we bought a new home and have been working on the flower garden in few more days we we done with the bambo wall n my garden we've really enjoyed the pertac | Severe | The speech was clear. Some words were combined or mispronounced. |
| bamboo walls a gan to be bery propular they are strong easy to use good looking to provide a good background and can e crate dok of a japanese garden bamboo is one of the largest most reveen growing grasses all over the world may varieties of bamboo boo a yar although it is also gon in amerigalast year bouht a new home and bwe moer flower bady fo more day we dumbed to bamboo bo my garden weve really enjoyed the barti | Severe | The speech was clear. Some words were combined or mispronounced or ended prematurely. There were multiple people speaking in the background due to the door being open during the recording. |
| bamboo walvs are getting to be very popular they are strong easy use and good looking they provide a good background and create the look of japanese garden bamboo is one of the largest and most rapidy be grass all f the world bany varieties of bamboo are grown in asia although it has also grown in america last year we had bought a new home and havea workingin thogarden in a few more days we resdoned with bamboo wall in our garden we have really enjoyed the mergant | Moderate | The speech was slurred. Some words were combined or mispronounced or ended prematurely. |
| now bamboo balls of gatin to be very popular they are strong easy use and good looking they provide good background and cancrate i look ofr jaminies o garden bamboo as woar the largest and most raputale ground graces all over the world many varieties of bamboo er gorn in asia up would also gor in america last year we brought im home an hapid were gol in a flower garden af yow more days we wwanowith bamboo ball n our garden we have really enjoyed the project | Moderate | The speech was slurred. Some words were combined or mispronounced or ended prematurely. |
| ban walls again ba baber they are strongesy dews an good looken e bavie ood background and gainade as a look of japanese grden of ad japanese garden bamboo is one of the largest and most gaaly growing grasses all over the world many drities of bamboo woard our given grond a natun all veris also grwn in amracker tus last year we bogd b new hoome an have mays we will be done with the bamboo wall and our gran we have we enjoyed the be barde | Severe | The speech was slurred and faint. Some words were combined or mispronounced or ended prematurely. |
| eobe barter are n te dels ean go eit ter oregebedgran nan a a rane o o are gaden e enes geraembors onenelodges a or aoote onn bsmet all r orer he ma raes oro enboror given ot ar aw an basure orn e oon dinins oof e orna rtar ter e ad the gadn tef the hee wo oon m aveworkeony ter rde donorgonin if yeu mr days we worgondeyol een we won be done a nbandbo als or garne we hav re enjon te broder | Severe | The speech was slurred and faint. There were multiple people speaking in the background due to the door being open during the recording. |
| bamblu halls are ca very to be very popular they are strong easuse and good looking be vride a be batron ad gan trat luck of dampis gardens bamblu is one of he largest and most rapidly bamgrasses all the world ba ver varieties of bamblu ar go n asia alloe and rome an america basoe got twoe houses barba and how have ben woking in the figarden a few more days and will becoe a bamblue hall ar frarget gardens we have fly enjoys of prospect | Severe | The speech was slurred, faint and almost unintelligible. |
| bamboo walls are getting to be very popular they are strong easy to use in good thuging they have providea garden bamboo is one of the largest most rapidly growing grasses all over the world mony fridays to bam boan our grown in asia although it is grown idimberga last year we've gogt a brand new home in have been worken on flower garden in a few more days we will be don with the bamboo wall in our garden we have really enjoyed the product | Moderate | The speech was slurred. The participant skipped the line *"good background and can create a look of a Japanese"* |
| bamboo walls a en berer ball or there are strong easy to use and good looking theyhave provided good byr ground and can greate locol japanese garden bamboos al the larges most rapidly gon grassis all the world many varieties of an bamboo at are gron in asia although wars alsso grown in america last year we bought a new home and ihave wred lon flower garden and a few moredays will be done with bamboo ball in our garden wee hav really enjoed the bardrarden | Severe | The speech was slurred, faint and almost unintelligible. Some words were combined, mispronounced or ended prematurely. |
| bamboo walls erkettinwe ery potbe art they are strong easy cus and good looking they brovid bood bad bo an gin greead of loo of ta japanese gardenn bamboo is wne bthe largest and most rapidly grwn grasses olover the worlld any vriyedays a bammboo werrgrown nasia wile though it is lso grown in amerigod last year wapwought a new home mhavboem boring on flowerard n in av few more daays weo betdown we bamboo balll in our gard w have vroienjoyed the project | Severe | The speech was faint and almost unintelligible. |
| bamboo balls are getting to be very popular they arststrong easy cuse and good looking they provided bood bad ground en gr at o ok of japanese garden bamboo is one of the largest most rapidly grown grasses all over the world any varieties o bamboo oure grown dasu all thotes also grow in america last year we bought a new ome and have been working g on the far gardens and af yew more days will be done with bamboo wall in our garden weave rillaendad the proget | Severe | The speech was faint and almost unintelligible. |
| bamboo wals are getting ery popular they're strong easy to use and good looking they provied a good back gon neand great look of a gjapanese garden bamboo is wone of the largest and mmost rapidly rowing grasses a lower ord many varieties o bamboo were grown in asia onthotis also rown in america last year we bought a new hhome and have been brking on the flower garden the fembor dis we te bitton with the bamboo ball and our garden we're bratly enjoyed the project | Severe | The speech was slurred and faint. Some words were not pronounced. |
| webore e embo golls ar gan to be aoga te ars rowing eservews in goo booking e oviede a wit barrarden a gree goevenen garde wbewoays won the rages an bos revre by growing grs aloe wold many varieties or be wor worinata abo it is alls o brwni in the margond lie here we brought ando won and hav een working on the flower aden an a few mrore days wen e regemrerebo all an nor garden we have really enjoyed the project | Severe | The speech was slurred, faint and almost unintelligible. Some words were combined or mispronounced or ended prematurely. There were multiple people speaking in the background due to the door being open during the recording. |
| s in e wond fnan tobe buri gbi mea strong a bines an gebnnin ti be fin bebbon in gin cree elo of d bmein its goninin as mn bonbes n mo srabo b groin grassens al bn the wold man a minn as a ben bel are rownen don all ovnas ovs ve grownin mrna las ai ben bin and in ha b boin on mod low gurnin imbod bated w ill be bnmn bin e bong in orn non we have winin i the bron migt | Severe | The speech was slurred, faint and almost unintelligible. Most words were combined or mispronounced or ended prematurely. There were multiple people speaking in the background due to the door being open during the recording. |
| who sa eitobeer botteetbteonaes toins tiis teoteto tae tnta teatena e tet en ae taesteg ae ne ete te teo te tietennto go go i goie gi esoo te bo oi tan te oti egaitise ei an i ii sen oo his is sow bon toea i ergan lest she be botton in n an ola anardan bo e tota garden if yeu more dais e o i ton n the an e e oin an d gand gadnein aand joid te drawjarkton | Moderate | The speech was faint and most words are unintelligible. |
| mambowalls are getting to be very popular they are strong isus and go in bambrading gon bambo and and grrade enew a look of ba japanese gardn bamboo deans bn of the largest and most rapidly rosgrasses all o erthe world many variations of bamboo ae rani esi although it is also brown nan a bar gelastyear wo oadani bo om e have been workin of the flowuer gardens in of newboar dads wo bedn a win the bamboo wall in ar gardenn we have really enjoyed the project | Moderate | The speech was slurred, faint and almost unintelligible. Some words were combined. |
| bamble walls are geting thed be very holpyo they are songy seals and good loking te provided to backgon and create elok of a japanese garden bambeg is one of gthe largest mossrably growing grass all over the world may we fridres of vamblu are gowing in asia although wit gov the groves in america larg year te pot i new home and have been working on the flower garden in a few more days we will be dn with the bamp wall in our garden we have really | Moderate | The speech was slurred, faint and almost unintelligible. Some words were combined. The recording did not include the final words *"enjoyed the project"* |
| bamboo walls ak gettin to be very populan they are strong easy to use in good looking tey proviede a good bapand and gcan treade the look ofjapanese garden bamboo is won the largest and most rapidbty grwng grates allob h world any varieties bthn bo a woimaga although ites te growin amaricalast year we brought a new home and have been working on the flower gardn an a few moretes woon by town of the bamboo pall nin our garden we have really enjoyed the frud | Moderate | The speech was slurred. Some words were combined. |
| bambovalls ar giving to be very populr gheerare srast eal easy to use and good looking the are brba go bacgrand and can craye it look for japanese garden bambois bonnop the largest and the bbost rrapidly growing grasses all over the w many erade abambo r or asia alto ed also going in america last yeavry bode ne bo and bi working an the flrar garden in fiol das be will be anui the banbobal at the garden we hav we rily enjored th bardy | Severe | The speech was slurred. Some words were combined or skipped. |
| bamboo walls like catic to wee very poplar they are strong easy to use and good looki bey boit a good becular encate to go japnese garden bamboo is ratof the larga pos ar petty gryy grasses ono the word bmany a litle bamboo ar roard in asia although it is also grown in america alte reportin o on i have been working on the flowr garden in a few more days will be done with the bamboo wall in a garden we have livi in your te prelic | Moderate | The speech was slurred. Some words were combined. |
| am wof igetet wi renaansayouth n golond i go vin i ot ba gan an crea d loot a a tobenies a bamo ats one of the liters an lat av oly owin gasen all over the world anly varieties af bambo ar goenin aga al to ate aso on an te ma a ar we wat ne o and hav en aamadewor gardn in s youod at we w we an wi am o world in or garden we ave rely and it ot | Moderate | The speech was unintelligible. |
| ambe welss are getting berry oaa a bavers gound esteduse and good looking tber ved a good grrund an tan reed a gokof japanese garden tam bws on anres an mos rapiley owingrasses all over the gord many arieties o banbe are grown in asia althouough it is also grown in ameriga last year wabat a newhowm and hav ben working on the gargd in a ew more days will be bon wit the bamb waw in our garden wehaverote enjoyed the godract | Severe | The speech was slurred. Some words were combined or skipped. |

*Supplementary Table 5 Example included and excluded transcripts along with the participant severity and description of the audio file*

#### Supplementary Methods 4. Learning the Acoustic-Linguistic Index of Ataxic Speech (ALIAS)

Least absolute shrinkage and selection operator (LASSO) regression was employed to derive the Acoustic-Linguistic Index for Ataxic Speech (ALIAS) that models the relationship between the extracted features and the BARS total score. As shown in Supplementary Table 6, separate ALIAS models were generated using different subsets of features. Model training and evaluation were conducted using leave-one-out cross-validation (LOOCV), in which all sessions from a single participant (including longitudinal recordings) were withheld as the test set, while data from the remaining participants were used for training. Within each LOOCV iteration, data normalization and hyperparameter optimization were performed independently. The regularization strength parameter was sampled 100 times from a log-uniform distribution between $10^{-3}$ and $10^{3}$, and the optimal value was determined via five-fold cross-validation within the training set. The final model was then retrained on the complete training data using the selected regularization parameter.

| Features | MFCC, μ | MFCC, σ | MFCC, μ and σ | μ MFCC σ | | Linguistic + MFCC σ | Linguistic + μ MFCC σ | All features |
|---|---|---|---|---|---|---|---|---|
| First four MFCCs (and their derivatives) | 0.27 | 0.63 | 0.59 | | - | 0.68 | | |
| All MFCCs (and their derivatives) | 0.20 | 0.59 | 0.57 | 0.62 | - | 0.66 | 0.68 | |
| Linguistic Only | | | | | 0.66 | | | 0.68 |

*Supplementary Table 6 Different ALIAS models and their correlation with BARS total score*

Training the ALIAS model using only the mean values of the mel-frequency cepstral coefficients (MFCCs) and their derivatives yielded suboptimal performance in predicting the total BARS score. In contrast, incorporating the standard deviation values of the MFCCs (and their derivatives) substantially improved predictive performance, suggesting that vocal instability – as captured by these variability measures – provides clinically informative cues for estimating disease severity. Combining both the mean and standard deviation values of MFCCs resulted in a reduced performance. This difference is likely driven by the relatively weak univariate correlations between mean values and the BARS total score, compared to the stronger correlations observed with the standard deviation features. As previously discussed in Supplementary Methods 2, an aggregate measure, μ MFCC σ, was computed by averaging the standard deviation values across MFCCs to account for their high intercorrelation. ALIAS models trained using either the first four MFCCs or all MFCCs demonstrated comparable performance. The ALIAS model based solely on linguistic features outperformed the MFCC-based model, and incorporating μ MFCC σ with linguistic features further enhanced predictive accuracy. Ultimately, the ALIAS model trained using all available acoustic and linguistic features exhibited the strongest association with the BARS total score.

| | Cross-Sectional (Controls, N=169) | | | | | | Longitudinal | | | | | | | | | | | | | |
|---|---|---|---|---|---|---|---|---|---|---|---|---|---|---|---|---|---|---|---|---|
| Features | vs Ataxic (N=223) | | vs without OSA (N=71) | | vs ataxic (N=30) | | Progression (N=97) | | Δ in Controls (N=43) | | Δ in Ataxia (N=54) | | Progression (N=56) | | Δ in individuals without OSA (N=13) | | Progression (N=48) | | Δ in pre-ataxic (N=5) | |
| | p | d | p | d | p | d | p | d | p | MSDR | p | MSDR | p | d | p | MSDR | p | d | p | MSDR |
| MFCC, μ | ** | -0.33 | | | | | | | | | | | | | | | | | | |
| MFCC, μ (first 4) | *** | -0.44 | | | | | | | | | | | | | | | | | | |
| MFCC, σ | *** | -1.34 | *** | -0.66 | * | -0.35 | | | | | | | | | | | | | | |
| MFCC, σ (first 4) | *** | -1.43 | *** | -0.72 | * | -0.45 | | | | | | | | | | | | | | |
| MFCC, μ and σ | *** | -1.32 | *** | -0.62 | | | | | | | | | * | 0.64 | * | -0.7 | * | 0.89 | | |
| MFCC, μ and σ (first 4) | *** | -1.39 | *** | -0.67 | * | -0.46 | | | | | | | | | | | | | | |
| μ MFCC σ | *** | -1.40 | *** | -0.68 | | | | | | | | | | | | | | | | |
| μ MFCC σ (first 4) | *** | -1.42 | *** | -0.70 | * | -0.37 | | | | | | | | | | | | | | |
| linguistic | *** | -1.12 | ** | -0.34 | | | | | | | | | | | | | | | | |
| Linguistic + MFCC σ | *** | -1.22 | *** | -0.43 | | | | | | | | | | | | | | | | |
| Linguistic + MFCC σ (first 4) | *** | -1.28 | *** | -0.50 | | | | | | | | | | | | | | | | |
| Linguistic + μ MFCC σ | *** | -1.33 | *** | -0.55 | | | | | | | | | | | | | | | | |
| Linguistic + μ MFCC σ (first 4) | *** | -1.32 | *** | -0.55 | | | | | | | | | | | | | | | | |
| ALL features | *** | -1.27 | *** | -0.48 | | | | | | | | | | | | | | | | |

| Feature | BARS | | | SARA | | MICARS | | | PROM ataxia | | | | DIS | CPIB |
|---|---|---|---|---|---|---|---|---|---|---|---|---|---|---|
| | Speech | Oculomotor | Total | Speech | Total | Fluency | Clarity | Alternating | Speech | Motor | Comm. | Total | Total | Total |
| MFCC, μ | 0.23 *** | 0.05 | 0.20 ** | 0.22 ** | 0.24 *** | 0.10 | 0.19 ** | 0.18 ** | 0.16 * | 0.16 * | 0.16 * | 0.16 * | -0.17 * | -0.09 |
| MFCC, μ (first 4) | 0.31 *** | 0.07 | 0.27 *** | 0.30 *** | 0.32 *** | 0.20 ** | 0.31 *** | 0.31 *** | 0.24 *** | 0.21 ** | 0.24 *** | 0.25 *** | -0.19 ** | -0.14 |
| MFCC, σ | 0.66 *** | 0.29 *** | 0.59 *** | 0.64 *** | 0.59 *** | 0.59 *** | 0.62 *** | 0.61 *** | 0.56 *** | 0.50 *** | 0.49 *** | 0.50 *** | -0.55 *** | -0.50 *** |
| MFCC, σ (first 4) | 0.69 *** | 0.31 *** | 0.63 *** | 0.66 *** | 0.63 *** | 0.61 *** | 0.64 *** | 0.61 *** | 0.56 *** | 0.51 *** | 0.48 *** | 0.51 *** | -0.56 *** | -0.52 *** |
| MFCC, μ and σ | 0.64 *** | 0.27 *** | 0.57 *** | 0.62 *** | 0.57 *** | 0.57 *** | 0.60 *** | 0.57 *** | 0.52 *** | 0.50 *** | 0.46 *** | 0.48 *** | -0.53 *** | -0.52 *** |
| MFCC, μ and σ (first 4) | 0.67 *** | 0.28 *** | 0.59 *** | 0.63 *** | 0.59 *** | 0.58 *** | 0.63 *** | 0.60 *** | 0.55 *** | 0.48 *** | 0.47 *** | 0.48 *** | -0.55 *** | -0.50 *** |
| μ MFCC σ | 0.66 *** | 0.31 *** | 0.62 *** | 0.63 *** | 0.61 *** | 0.57 *** | 0.60 *** | 0.61 *** | 0.57 *** | 0.51 *** | 0.50 *** | 0.50 *** | -0.58 *** | -0.55 *** |
| μ MFCC σ (first 4) | 0.68 *** | 0.29 *** | 0.63 *** | 0.64 *** | 0.64 *** | 0.58 *** | 0.62 *** | 0.61 *** | 0.59 *** | 0.53 *** | 0.49 *** | 0.52 *** | -0.58 *** | -0.55 *** |
| linguistic | 0.67 *** | 0.23 *** | 0.66 *** | 0.62 *** | 0.67 *** | 0.61 *** | 0.62 *** | 0.59 *** | 0.50 *** | 0.49 *** | 0.44 *** | 0.46 *** | -0.57 *** | -0.49 *** |
| Linguistic + MFCC σ | 0.68 *** | 0.26 *** | 0.66 *** | 0.65 *** | 0.66 *** | 0.62 *** | 0.64 *** | 0.63 *** | 0.58 *** | 0.52 *** | 0.50 *** | 0.51 *** | -0.62 *** | -0.54 *** |
| Linguistic + MFCC σ (first 4) | 0.70 *** | 0.29 *** | 0.68 *** | 0.66 *** | 0.68 *** | 0.63 *** | 0.64 *** | 0.61 *** | 0.55 *** | 0.53 *** | 0.48 *** | 0.50 *** | -0.60 *** | -0.53 *** |
| Linguistic + μ MFCC σ | 0.69 *** | 0.29 *** | 0.68 *** | 0.65 *** | 0.67 *** | 0.62 *** | 0.63 *** | 0.61 *** | 0.59 *** | 0.54 *** | 0.52 *** | 0.53 *** | -0.64 *** | -0.56 *** |
| Linguistic + μ MFCC σ (first 4) | 0.69 *** | 0.29 *** | 0.68 *** | 0.65 *** | 0.68 *** | 0.61 *** | 0.63 *** | 0.60 *** | 0.58 *** | 0.54 *** | 0.50 *** | 0.52 *** | -0.62 *** | -0.56 *** |
| ALL features | 0.68 *** | 0.25 *** | 0.68 *** | 0.65 *** | 0.67 *** | 0.62 *** | 0.66 *** | 0.63 *** | 0.53 *** | 0.51 *** | 0.48 *** | 0.50 *** | -0.58 *** | -0.49 *** |

#### Supplementary Methods 5. Different Sampling Strategies for Reliability Analysis

To compute the robustness and consistency of the speech-derived features, we calculated reliability using three distinct sampling strategies designed to partition each recorded session into equal segments. These methods allowed us to evaluate whether acoustic and linguistic markers remain stable across different portions of a single task or when subjected to random variation:

- $\mathcal{R}_1$ (temporal split): The voiced session was divided into two chronological halves—the first half and the second half. This strategy allows us to assess the stability of features over the duration of the task.
- $\mathcal{R}_2$ (randomized split): We extracted two random halves from the voiced session. By sampling non-contiguous segments, this strategy allows us to evaluate the influence of temporal trends in the consistency of the acoustic feature.
- $\mathcal{R}_3$ (word-level shuffle): We shuffled the spoken words within the session before extracting two equal halves. This strategy controls for the specific phonetic context of the passage, ensuring that the extracted features are inherent to the speaker's motor execution rather than being biased by the specific sequence of words.

By employing these varied strategies, we ensure that the extracted speech measures are not artifacts of specific sampling windows but are reliable indicators of the speaker’s speech characteristics.

| Feature Name | $\mathcal{R}_1$ | $\mathcal{R}_2$ | $\mathcal{R}_3$ | Feature Name | $\mathcal{R}_1$ | $\mathcal{R}_2$ | $\mathcal{R}_3$ | Feature Name | $\mathcal{R}_1$ | $\mathcal{R}_2$ | $\mathcal{R}_3$ |
|---|---|---|---|---|---|---|---|---|---|---|---|
| MFCC1, μ | 0.96 | 1 | 0.98 | MFCC1’, μ | -0.87 | -0.08 | 0.08 | MFCC1’’, μ | -0.22 | -0.07 | 0.07 |
| MFCC2, μ | 0.77 | 0.99 | 0.92 | MFCC2’, μ | -0.76 | 0.02 | 0 | MFCC2’’, μ | -0.14 | 0.02 | 0.08 |
| MFCC3, μ | 0.96 | 1 | 0.95 | MFCC3’, μ | -0.83 | -0.01 | 0.02 | MFCC3’’, μ | -0.12 | 0.06 | 0.06 |
| MFCC4, μ | 0.94 | 1 | 0.9 | MFCC4’, μ | -0.87 | 0.04 | -0.05 | MFCC4’’, μ | -0.12 | 0.01 | 0.13 |
| MFCC5, μ | 0.97 | 1 | 0.95 | MFCC5’, μ | -0.84 | 0.01 | 0.04 | MFCC5’’, μ | -0.01 | -0.02 | 0.07 |
| MFCC6, μ | 0.94 | 1 | 0.96 | MFCC6’, μ | -0.82 | 0.06 | 0.09 | MFCC6’’, μ | -0.01 | 0.04 | 0 |
| MFCC7, μ | 0.96 | 1 | 0.97 | MFCC7’, μ | -0.72 | 0.05 | -0.07 | MFCC7’’, μ | 0.03 | 0 | 0.02 |
| MFCC8, μ | 0.96 | 1 | 0.98 | MFCC8’, μ | -0.73 | -0.08 | -0.1 | MFCC8’’, μ | 0.01 | 0.03 | -0.02 |
| MFCC9, μ | 0.95 | 1 | 0.96 | MFCC9’, μ | -0.73 | -0.07 | -0.08 | MFCC9’’, μ | 0.08 | -0.08 | 0.03 |
| MFCC10, μ | 0.96 | 1 | 0.97 | MFCC10’, μ | -0.7 | 0.06 | 0.02 | MFCC10’’, μ | 0.06 | -0.01 | 0.05 |
| MFCC11, μ | 0.95 | 1 | 0.96 | MFCC11’, μ | -0.67 | -0.11 | -0.05 | MFCC11’’, μ | 0.06 | -0.05 | 0.04 |
| MFCC12, μ | 0.95 | 1 | 0.97 | MFCC12’, μ | -0.56 | 0.06 | 0.04 | MFCC12’’, μ | 0.12 | 0.07 | 0.04 |
| MFCC13, μ | 0.95 | 1 | 0.97 | MFCC13’, μ | -0.65 | -0.02 | 0.03 | MFCC13’’, μ | -0.03 | -0.02 | 0.02 |
| MFCC1, σ | 0.92 | 0.99 | 0.91 | MFCC1’, σ | 0.88 | 0.99 | 0.92 | MFCC1’’, σ | 0.86 | 0.99 | 0.87 |
| MFCC2, σ | 0.84 | 0.97 | 0.77 | MFCC2’, σ | 0.65 | 0.97 | 0.75 | MFCC2’’, σ | 0.5 | 0.96 | 0.62 |
| MFCC3, σ | 0.87 | 0.99 | 0.93 | MFCC3’, σ | 0.9 | 0.99 | 0.95 | MFCC3’’, σ | 0.88 | 0.99 | 0.9 |
| MFCC4, σ | 0.86 | 0.99 | 0.87 | MFCC4’, σ | 0.8 | 0.98 | 0.8 | MFCC4’’, σ | 0.64 | 0.98 | 0.67 |
| MFCC5, σ | 0.9 | 0.99 | 0.81 | MFCC5’, σ | 0.84 | 0.98 | 0.84 | MFCC5’’, σ | 0.69 | 0.98 | 0.72 |
| MFCC6, σ | 0.86 | 0.98 | 0.88 | MFCC6’, σ | 0.7 | 0.98 | 0.81 | MFCC6’’, σ | 0.49 | 0.97 | 0.64 |
| MFCC7, σ | 0.85 | 0.98 | 0.85 | MFCC7’, σ | 0.81 | 0.97 | 0.8 | MFCC7’’, σ | 0.63 | 0.97 | 0.63 |
| MFCC8, σ | 0.85 | 0.98 | 0.89 | MFCC8’, σ | 0.77 | 0.98 | 0.78 | MFCC8’’, σ | 0.6 | 0.97 | 0.62 |
| MFCC9, σ | 0.85 | 0.98 | 0.85 | MFCC9’, σ | 0.73 | 0.97 | 0.71 | MFCC9’’, σ | 0.58 | 0.96 | 0.58 |
| MFCC10, σ | 0.86 | 0.98 | 0.88 | MFCC10’, σ | 0.71 | 0.98 | 0.74 | MFCC10’’, σ | 0.6 | 0.97 | 0.6 |
| MFCC11, σ | 0.82 | 0.97 | 0.85 | MFCC11’, σ | 0.64 | 0.96 | 0.66 | MFCC11’’, σ | 0.56 | 0.95 | 0.55 |
| MFCC12, σ | 0.83 | 0.97 | 0.83 | MFCC12’, σ | 0.73 | 0.96 | 0.67 | MFCC12’’, σ | 0.67 | 0.95 | 0.62 |
| MFCC13, σ | 0.81 | 0.97 | 0.83 | MFCC13’, σ | 0.71 | 0.96 | 0.7 | MFCC13’’, σ | 0.65 | 0.95 | 0.61 |
| Mean MFCC σ | 0.95 | 1.00 | 0.97 | Mean MFCC’ σ | 0.87 | 1.0 | 0.94 | Mean MFCC’’ σ | 0.82 | 0.99 | 0.88 |
| chroma1, μ | 0.90 | 0.99 | 0.93 | chroma1’, μ | -0.61 | 0.04 | 0.03 | chroma1’’, μ | 0.15 | -0.02 | -0.03 |
| chroma2, μ | 0.90 | 0.99 | 0.94 | chroma2’, μ | -0.66 | -0.03 | 0.02 | chroma2’’, μ | 0.13 | 0.00 | 0.00 |
| chroma3, μ | 0.90 | 1.00 | 0.95 | chroma3’, μ | -0.67 | -0.01 | 0.02 | chroma3’’, μ | 0.13 | 0.01 | 0.02 |
| chroma4, μ | 0.92 | 1.00 | 0.95 | chroma4’, μ | -0.64 | 0.02 | 0.03 | chroma4’’, μ | 0.09 | 0.00 | 0.03 |
| chroma5, μ | 0.91 | 0.99 | 0.95 | chroma5’, μ | -0.67 | 0.02 | 0.11 | chroma5’’, μ | 0.07 | 0.03 | -0.03 |
| chroma6, μ | 0.90 | 0.99 | 0.94 | chroma6’, μ | -0.73 | -0.06 | 0.11 | chroma6’’, μ | -0.03 | 0.00 | -0.05 |
| chroma7, μ | 0.89 | 0.99 | 0.93 | chroma7’, μ | -0.73 | -0.01 | -0.04 | chroma7’’, μ | -0.02 | -0.05 | -0.11 |
| chroma8, μ | 0.86 | 0.99 | 0.92 | chroma8’, μ | -0.77 | 0.02 | 0.05 | chroma8’’, μ | -0.01 | -0.05 | -0.08 |
| chroma9, μ | 0.89 | 0.99 | 0.93 | chroma9’, μ | -0.74 | 0.07 | 0.01 | chroma9’’, μ | 0.06 | -0.05 | -0.02 |
| chroma10, μ | 0.90 | 0.99 | 0.94 | chroma10’, μ | -0.66 | 0.08 | 0.03 | chroma10’’, μ | 0.05 | -0.04 | -0.04 |
| chroma11, μ | 0.89 | 0.99 | 0.92 | chroma11’, μ | -0.63 | -0.05 | 0.03 | chroma11’’, μ | 0.07 | -0.02 | -0.04 |
| chroma12, μ | 0.89 | 0.99 | 0.93 | chroma12’, μ | -0.59 | 0.01 | 0.01 | chroma12’’, μ | 0.12 | -0.01 | -0.02 |
| chroma1, σ | 0.79 | 0.97 | 0.84 | chroma1’, σ | 0.75 | 0.96 | 0.85 | chroma1’’, σ | 0.76 | 0.95 | 0.83 |
| chroma2, σ | 0.79 | 0.97 | 0.86 | chroma2’, σ | 0.77 | 0.96 | 0.86 | chroma2’’, σ | 0.77 | 0.95 | 0.85 |
| chroma3, σ | 0.76 | 0.97 | 0.82 | chroma3’, σ | 0.75 | 0.94 | 0.84 | chroma3’’, σ | 0.75 | 0.92 | 0.82 |
| chroma4, σ | 0.81 | 0.97 | 0.80 | chroma4’, σ | 0.77 | 0.94 | 0.85 | chroma4’’, σ | 0.78 | 0.91 | 0.84 |
| chroma5, σ | 0.86 | 0.98 | 0.89 | chroma5’, σ | 0.80 | 0.96 | 0.85 | chroma5’’, σ | 0.80 | 0.95 | 0.83 |
| chroma6, σ | 0.87 | 0.99 | 0.91 | chroma6’, σ | 0.81 | 0.97 | 0.85 | chroma6’’, σ | 0.81 | 0.95 | 0.83 |
| chroma7, σ | 0.89 | 0.99 | 0.90 | chroma7’, σ | 0.82 | 0.97 | 0.87 | chroma7’’, σ | 0.86 | 0.95 | 0.88 |
| chroma8, σ | 0.86 | 0.98 | 0.89 | chroma8’, σ | 0.81 | 0.97 | 0.86 | chroma8’’, σ | 0.85 | 0.95 | 0.87 |
| chroma9, σ | 0.81 | 0.98 | 0.87 | chroma9’, σ | 0.76 | 0.96 | 0.85 | chroma9’’, σ | 0.78 | 0.94 | 0.85 |
| chroma10, σ | 0.82 | 0.98 | 0.87 | chroma10’, σ | 0.76 | 0.96 | 0.85 | chroma10’’, σ | 0.78 | 0.95 | 0.85 |
| chroma11, σ | 0.78 | 0.97 | 0.82 | chroma11’, σ | 0.80 | 0.94 | 0.86 | chroma11’’, σ | 0.79 | 0.93 | 0.84 |
| chroma12, σ | 0.75 | 0.96 | 0.83 | chroma12’, σ | 0.77 | 0.94 | 0.84 | chroma12’’, σ | 0.78 | 0.93 | 0.83 |
| μ chroma σ | 0.88 | 0.99 | 0.93 | μ chroma’ σ | 0.79 | 0.98 | 0.88 | μ chroma’’ σ | 0.81 | 0.97 | 0.88 |
| slope, μ | 0.97 | 1.00 | 0.99 | slope', μ | -0.96 | 0.03 | 0.01 | slope", μ | 0.00 | -0.05 | -0.09 |
| slope, σ | 0.94 | 1.00 | 0.97 | slope', σ | 0.94 | 1.00 | 0.97 | slope", σ | 0.92 | 1.00 | 0.95 |
| flux, μ | 0.97 | 1.00 | 0.98 | flux', μ | -0.72 | 0.03 | -0.10 | flux", μ | 0.11 | -0.17 | 0.02 |
| flux, σ | 0.74 | 0.98 | 0.78 | flux', σ | 0.77 | 0.98 | 0.81 | flux", σ | 0.73 | 0.98 | 0.77 |
| entropy, μ | 0.86 | 0.99 | 0.95 | entropy', μ | -0.45 | 0.10 | -0.04 | entropy",μ | -0.12 | -0.13 | -0.01 |
| entropy, σ | 0.89 | 0.99 | 0.88 | entropy', σ | 0.86 | 0.98 | 0.87 | entropy", σ | 0.83 | 0.98 | 0.81 |

| | | | | | | | | | | | |
|---|---|---|---|---|---|---|---|---|---|---|---|
| centroid,μ | 0.70 | 0.99 | 0.86 | centroid', μ | -0.42 | 0.09 | 0.00 | centroid", μ | -0.15 | -0.05 | 0.07 |
| centroid, σ | 0.57 | 0.95 | 0.61 | centroid', σ | 0.58 | 0.95 | 0.73 | centroid", σ | 0.52 | 0.94 | 0.67 |
| spread, μ | 0.67 | 0.99 | 0.90 | spread', μ | -0.72 | -0.02 | -0.01 | spread", μ | -0.09 | 0.00 | 0.01 |
| spread, σ | 0.48 | 0.96 | 0.67 | spread', σ | 0.60 | 0.95 | 0.81 | spread", σ | 0.55 | 0.94 | 0.74 |
| skewness, μ | 0.79 | 0.99 | 0.94 | skewness', μ | -0.63 | -0.05 | 0.00 | skewness", μ | -0.16 | -0.01 | -0.09 |
| skewness, σ | 0.91 | 0.99 | 0.88 | skewness', σ | 0.91 | 0.98 | 0.90 | skewness", σ | 0.87 | 0.97 | 0.86 |
| kurtosis, μ | 0.79 | 1.00 | 0.94 | kurtosis', μ | -0.74 | 0.04 | -0.04 | kurtosis", μ | -0.08 | -0.05 | -0.02 |
| kurtosis, σ | 0.86 | 0.99 | 0.91 | kurtosis', σ | 0.89 | 0.99 | 0.93 | kurtosis", σ | 0.87 | 0.99 | 0.90 |
| flatness, μ | 0.61 | 0.97 | 0.74 | flatness', μ | -0.24 | 0.00 | 0.00 | flatness", μ | -0.02 | 0.02 | -0.01 |
| flatness, σ | 0.69 | 0.94 | 0.64 | flatness', σ | 0.65 | 0.94 | 0.66 | flatness", σ | 0.59 | 0.94 | 0.61 |
| rolloff, μ | 0.68 | 0.99 | 0.88 | rolloff', μ | -0.42 | -0.09 | 0.04 | rolloff", μ | -0.09 | 0.02 | 0.00 |
| rolloff, σ | 0.51 | 0.95 | 0.65 | rolloff', σ | 0.64 | 0.95 | 0.80 | rolloff", σ | 0.61 | 0.94 | 0.77 |
| rms, μ | 0.96 | 1.00 | 0.98 | rms', μ | -0.96 | -0.12 | -0.02 | rms", μ | -0.07 | -0.05 | 0.17 |
| rms, σ | 0.90 | 1.00 | 0.94 | rms', σ | 0.90 | 1.00 | 0.95 | rms", σ | 0.88 | 1.00 | 0.93 |
| crest_factor, μ | 0.84 | 0.97 | 0.94 | crest_factor', μ | -0.22 | 0.08 | 0.03 | crest_factor", μ | -0.04 | 0.06 | 0.10 |
| crest_factor, σ | 0.78 | 0.95 | 0.90 | crest_factor', σ | 0.80 | 0.95 | 0.88 | crest_factor", σ | 0.80 | 0.94 | 0.87 |
| f0_contour, μ | 0.97 | 1.00 | 0.97 | f0_contour', μ | -0.56 | 0.03 | -0.01 | f0_contour", μ | -0.05 | -0.08 | -0.12 |
| f0_contour, σ | 0.88 | 0.99 | 0.92 | f0_contour', σ | 0.81 | 0.99 | 0.89 | f0_contour", σ | 0.80 | 0.98 | 0.86 |
| kurtosis, μ | 0.82 | 0.98 | 0.92 | kurtosis', μ | -0.26 | -0.08 | -0.06 | kurtosis", μ | 0.01 | 0.10 | 0.01 |
| kurtosis, σ | 0.70 | 0.89 | 0.80 | kurtosis', σ | 0.64 | 0.88 | 0.72 | kurtosis", σ | 0.65 | 0.88 | 0.70 |
| Shannon E, μ | 0.97 | 1.00 | 0.98 | Shannon E', μ | -0.92 | -0.07 | 0.05 | Shannon E", μ | -0.29 | -0.31 | 0.44 |
| Shannon E, σ | 0.88 | 0.99 | 0.90 | Shannon E', σ | 0.81 | 0.99 | 0.88 | Shannon E", σ | 0.77 | 0.99 | 0.85 |
| Speaking Rate | 0.81 | | | | | | | | | | |
| PD, μ | 0.90 | | | | | | | | | | |
| PD, σ | 0.70 | | | | | | | | | | |
| Percent Pause | 0.73 | | | | | | | | | | |
| Pause Events | -0.39 | | | | | | | | | | |
| Speech Duration | 0.84 | | | | | | | | | | |
| Total Duration | 0.99 | | | | | | | | | | |
| Total PD | 0.96 | | | | | | | | | | |

*Supplementary Table 7 Reliability of speech-derived features across different sampling strategies* $\mathcal{R}_1: First\ Half, Second\ Half$ $\mathcal{R}_2: Random\ Halve$ $\mathcal{R}_3: Random\ Shuffle\ Words$

#### Supplementary Methods 6. Sustained Vowel Phonation Task

In this task, participants were asked to sustain a vowel (*/ah/*) at their normal speaking volume. Before they begin, participants were told to take a deep breath and to continue the task for at least 10 seconds. They were also instructed to keep an even tone and try not to sing. This task was intended to measure fluctuations in frequency and amplitude and vocal alterations during the task. The sustained vowel phonation sessions completed by the same participants that were included in the analysis of the passage reading task were analyzed.

In our passage reading task analyses, different measures of μ MFCC σ were effective in distinguishing individuals with ataxia from healthy controls, identifying ataxic individuals without OSA and capturing longitudinal alterations in speech patterns associated with disease progression. To further investigate this measure, we extracted μ MFCC σ features (and its derivatives) during a sustained vowel phonation (*/ah/*) task performed by the same participants (More information in the Supplementary Methods 5). We found that μ MFCC σ (and its derivatives) were also effective in distinguishing individuals with ataxia from healthy controls (μ MFCC σ: $p<0.001$, d=-0.46, μ MFCC' σ: $p<0.001$, d=-0.41 and μ MFCC'' σ: $p<0.001$, d=-0.35) and identifying ataxic individuals with no dysarthria (μ MFCC' σ: $p<0.05$, d=-0.31 and μ MFCC'' σ: $p<0.05$, d=-0.27). However, only μ MFCC σ was able to capture longitudinal alterations in speech patterns associated with disease progression in the ataxic population ($p<0.01$, MSDR=-0.32).

### Supplementary Results

*Supplementary Table 8 The mean (μ) and standard deviation (σ) of different MFCC extracted from the voiced segments during passage reading. The construct validity of the extracted feature was assessed through: (1) comparison between Ataxias and Control, (2) early detection between ataxia individuals with BARS speech = 0 and Controls, (3) Disease Progression Comparison between Ataxias and Control and (4) Analysis of Longitudinal Change with the Ataxia Population. All results include the p-value (Cohen's d value). <0.05*, <0.01**, <0.001****

| | Cross-Sectional | | | | | | Longitudinal | | | | | | | | | | | | | |
|---|---|---|---|---|---|---|---|---|---|---|---|---|---|---|---|---|---|---|---|---|
| Feature | vs Ataxic (N=395) | | vs NOSA (N=240) | | vs pre-ataxic (N=199) | | Progression (N=97) | | Δ in Controls (N=43) | | Δ in Ataxia (N=54) | | Progression (N=56) | | Δ in NOSA (N=13) | | Progression (N=48) | | Δ in preataxic (N=5) | |
| | p | d | p | d | p | d | p | d | p | MSDR | p | MSDR | p | d | p | MSDR | p | d | p | MSDR |
| MFCC1, μ | * | -0.45 | | | | | | | | | | | | | * | -0.64 | | | | |
| MFCC2, μ | | | * | -0.42 | | | | | ** | 0.41 | * | 0.48 | | | ** | 1.56 | | | * | 1.25 |
| MFCC3, μ | | | | | | | | | | | | | | | | | | | | |
| MFCC4, μ | | | | | | | | | * | -0.29 | * | -0.54 | | | | | | | | |
| MFCC5, μ | | | | | | | * | -0.47 | ** | -0.61 | | | | | ** | -1.34 | | | * | -4.77 |
| MFCC6, μ | | | | | * | -0.45 | * | -0.34 | * | -0.36 | | | | | | | | | | |
| MFCC7, μ | | | | | | | | | ** | 0.34 | | | | | ** | 1.37 | | | * | 1.94 |
| MFCC8, μ | | | | | ** | -0.64 | | | | | | | | | * | 0.84 | | | * | 1.40 |
| MFCC9, μ | | | * | 0.39 | | | | | * | 0.36 | | | | | * | 0.62 | | | | |
| MFCC10, μ | | | | | | | | | * | 0.45 | | | | | | | | | | |
| MFCC11, μ | | | | | | | | | * | 0.38 | | | | | * | 0.67 | | | | |
| MFCC12, μ | | | | | | | | | | | | | | | | | | | | |
| MFCC13, μ | | | | | | | | | | | | | ** | 0.66 | * | -1.12 | | | | |
| MFCC1, σ | | | | | | | | | | | * | -0.39 | | | | | | | | |
| MFCC2, σ | | | | | | | | | | | | | | | | | | | | |
| MFCC3, σ | | | * | 0.43 | | | | | | | * | -0.54 | | | | | | | | |
| MFCC4, σ | | | | | | | | | | | | | | | | | | | | |
| MFCC5, σ | | | | | | | | | | | | | | | | | | | | |
| MFCC6, σ | | | | | | | | | | | | | | | | | | | | |
| MFCC7, σ | | | | | | | | | | | * | -0.48 | | | | | | | | |
| MFCC8, σ | | | | | | | | | | | | | | | | | | | | |
| MFCC9, σ | | | | | | | | | | | | | | | | | | | | |
| MFCC10, σ | | | | | | | | | | | | | | | | | | | | |
| MFCC11, σ | | | | | | | ** | 0.37 | | | * | -0.44 | | | * | -0.82 | | | | |
| MFCC12, σ | | | | | | | | | | | | | ** | 0.71 | * | -1.01 | | | | |
| MFCC13, σ | * | -0.49 | * | -0.43 | | | | | | | | | | | | | | | | |
| μ MFCC σ | | | | | | | | | | | * | -0.41 | | | | | | | | |
| chroma1, μ | | | * | -0.57 | ** | -0.72 | | | | | | | | | | | | | | |
| chroma2, μ | | | * | -0.44 | ** | -0.68 | | | | | | | | | | | | | | |
| chroma3, μ | | | | | * | -0.63 | | | | | | | | | | | | | | |
| chroma4, μ | | | | | ** | -0.60 | | | | | | | | | | | | | | |
| chroma5, μ | | | | | ** | -0.62 | | | | | | | | | | | | | | |
| chroma6, μ | | | | | | | | | | | | | | | | | | | | |
| chroma7, μ | | | | | | | ** | -0.39 | * | -0.34 | | | | | | | ** | -0.73 | | |
| chroma8, μ | | | | | | | ** | -0.36 | * | -0.33 | | | | | | | ** | -0.57 | | |
| chroma9, μ | | | | | | | | | | | | | | | | | | | | |
| chroma10, μ | | | | | | | | | | | | | | | | | | | * | 1.76 |
| chroma11, μ | | | | | | | | | | | | | | | | | | | | |
| chroma12, μ | | | * | -0.43 | * | -0.43 | | | | | | | | | | | | | | |
| chroma1, σ | * | -0.46 | | | | | * | 0.47 | | | | | | | | | | | | |
| chroma2, σ | | | | | | | | | * | 0.46 | | | | | | | | | | |
| chroma3, σ | | | | | | | | | * | 0.32 | | | | | | | | | | |
| chroma4, σ | | | | | | | | | | | | | | | | | | | | |
| chroma5, σ | | | | | * | 0.45 | | | | | | | | | | | | | | |
| chroma6, σ | | | | | ** | 0.73 | * | 0.43 | | | | | | | | | | | | |
| chroma7, σ | | | * | 0.47 | ** | 0.72 | | | | | | | | | | | | | | |
| chroma8, σ | | | | | | | | | | | | | | | | | | | | |
| chroma9, σ | | | | | | | | | * | 0.37 | | | | | * | 0.70 | | | | |
| chroma10, σ | | | | | * | 0.48 | | | | | * | 0.34 | | | | | | | | |
| chroma11, σ | | | | | | | | | | | | | | | | | | | | |
| chroma12, σ | | | | | | | | | | | | | | | | | | | | |
| μ chroma σ | | | | | | | * | 0.38 | * | 0.32 | | | | | | | | | | |
| slope, m | * | 0.42 | | | | | | | * | 0.35 | * | 0.26 | | | | | | | * | 1.81 |
| slope, s | * | -0.46 | | | | | | | | | * | -0.31 | | | | | | | * | -1.44 |
| flux, m | | | | | | | | | | | * | -0.29 | | | | | | | * | -1.84 |
| flux, s | * | -0.37 | | | | | | | | | * | -0.29 | | | | | | | * | -2.41 |
| entropy, m | | | | | | | | | * | 0.29 | | | ** | 0.61 | | | | | | |
| entropy, s | | | | | | | | | ** | -0.58 | * | -0.67 | | | | | | | | |
| centroid,m | | | * | 0.32 | | | | | | | | | ** | 0.69 | ** | -1.17 | | | | |
| centroid, s | | | * | 0.47 | | | | | | | | | | | | | | | | |
| spread, m | | | * | 0.30 | | | | | | | | | | | * | -0.78 | | | | |
| spread, s | | | ** | 0.52 | * | 0.33 | | | | | | | | | | | | | * | 2.12 |
| skewness, m | | | * | -0.30 | | | | | | | | | * | -0.58 | ** | 1.01 | | | * | 1.02 |
| skewness, s | | | | | | | | | | | | | | | | | | | | |
| kurtosis, m | | | * | -0.30 | | | | | | | | | * | -0.51 | ** | 0.91 | | | | |
| kurtosis, s | | | | | | | | | | | | | | | | | | | | |
| flatness, m | | | | | | | | | | | | | | | * | -0.72 | | | | |
| flatness, s | | | | | | | | | | | | | | | | | | | | |
| rolloff, m | | | * | 0.35 | | | | | | | | | * | 0.62 | ** | -0.88 | | | | |
| rolloff, s | * | 0.43 | ** | 0.53 | * | 0.37 | | | | | | | | | | | | | | |
| rms, m | * | -0.43 | | | | | | | * | -0.45 | * | -0.30 | | | | | | | * | -2.11 |
| rms, s | * | -0.51 | | | | | | | | | * | -0.37 | | | | | | | * | -1.28 |
| crest_factor, m | | | | | | | | | | | | | | | | | | | | |
| crest_factor, s | | | | | | | | | | | ** | -0.49 | | | | | | | | |
| f0_contour, m | | | * | 0.39 | | | ** | -0.96 | ** | -0.92 | | | ** | -1.16 | | | ** | -1.19 | | |
| f0_contour, s | | | | | | | | | | | | | | | | | | | | |
| kurtosis, m | | | | | | | * | 0.56 | | | | | | | | | | | | |
| kurtosis, s | | | | | | | | | | | * | -0.44 | | | | | | | | |
| Shannon E, m | * | -0.34 | | | | | | | * | -0.39 | * | -0.29 | | | | | | | * | -1.81 |
| Shannon E, s | * | -0.39 | | | | | | | | | * | -0.34 | | | | | | | * | -1.24 |

| | | | | | | | | | | | | | | | | |
|---|---|---|---|---|---|---|---|---|---|---|---|---|---|---|---|---|
| Speaking Rate | ** | 1.23 | * | 0.40 | | | | | | | | | | | | |
| Pause Duration, μ | ** | -0.91 | ** | -0.35 | | | | | | | | | | | | |
| Pause Duration, σ | ** | -0.64 | * | -0.37 | | | | | | | | | | | | |
| Percent Pause | ** | -0.87 | * | -0.47 | | | | | | | | | | | | |
| Pause Events | | | | | | | | | | | | | | | * | -1.19 |
| Speech Duration | ** | -1.01 | * | -0.29 | | | | | | | | | | | | |
| Total Duration | ** | -0.95 | * | -0.32 | | | | | | | | | | | | |
| Total Pause Duration | ** | -0.86 | ** | -0.35 | | | | | | | | | | | | |

| | Cross-Sectional | | | | | | Longitudinal | | | | | | | | | | | | | |
|---|---|---|---|---|---|---|---|---|---|---|---|---|---|---|---|---|---|---|---|---|
| Feature | vs Ataxic (N=395) | | vs NOSA (N=240) | | vs pre-ataxic (N=199) | | Progression (N=97) | | Δ in Controls (N=43) | | Δ in Ataxia (N=54) | | Progression (N=56) | | Δ in NOSA (N=13) | | Progression (N=48) | | Δ in preataxic (N=5) | |
| | p | d | p | d | p | d | p | d | p | MSDR | p | MSDR | p | d | p | MSDR | p | d | p | MSDR |
| MFCC1’, μ | | | | | | | | | | | | | | | | | | | | |
| MFCC2’, μ | | | | | | | | | | | | | | | | | | | | |
| MFCC3’, μ | | | | | | | | | | | | | | | | | | | * | 2.59 |
| MFCC4’, μ | | | | | | | | | | | | | | | | | | | | |
| MFCC5’, μ | | | | | | | | | | | | | | | | | | | | |
| MFCC6’, μ | | | | | | | | | | | | | | | | | | | | |
| MFCC7’, μ | | | | | | | | | | | | | | | | | | | | |
| MFCC8’, μ | | | | | | | | | | | | | | | | | | | | |
| MFCC9’, μ | | | | | | | | | | | | | | | | | | | | |
| MFCC10’, μ | | | | | | | | | | | | | | | | | | | | |
| MFCC11’, μ | | | | | | | | | | | | | | | | | | | | |
| MFCC12’, μ | | | | | | | | | | | | | | | | | | | | |
| MFCC13’, μ | | | | | | | | | | | | | | | | | | | | |
| MFCC1’, σ | | | * | 0.38 | | | | | | | * | -0.49 | | | | | | | | |
| MFCC2’, σ | * | 0.67 | * | 0.30 | | | | | | | * | -0.47 | | | | | | | | |
| MFCC3’, σ | * | 0.65 | ** | 0.56 | * | 0.45 | | | | | ** | -0.75 | | | | | | | | |
| MFCC4’, σ | * | 0.61 | * | 0.35 | | | | | | | * | -0.31 | | | | | | | | |
| MFCC5’, σ | | | | | | | | | * | 0.35 | | | | | ** | 1.08 | | | * | 1.54 |
| MFCC6’, σ | * | 0.45 | | | * | 0.39 | * | 0.38 | | | | | | | | | ** | 0.58 | | |
| MFCC7’, σ | * | 0.69 | | | | | | | | | | | | | | | | | | |
| MFCC8’, σ | * | 0.54 | | | * | 0.44 | | | | | | | | | | | | | | |
| MFCC9’, σ | * | 0.52 | | | | | | | | | | | | | | | | | | |
| MFCC10’, σ | * | 0.51 | | | | | | | | | | | | | | | | | | |
| MFCC11’, σ | * | 0.51 | | | | | | | | | | | | | | | | | | |
| MFCC12’, σ | | | | | | | | | | | | | | | | | | | | |
| MFCC13’, σ | | | | | | | | | | | | | | | | | | | | |
| μMFCC’σ | * | 0.64 | * | 0.41 | | | | | | | * | -0.57 | | | | | | | | |
| chroma1’, μ | | | | | | | | | | | | | | | | | | | | |
| chroma2’, μ | | | | | | | | | | | | | | | | | | | | |
| chroma3’, μ | | | | | | | | | | | | | | | | | | | | |
| chroma4’, μ | | | | | | | | | | | | | | | | | | | | |
| chroma5’, μ | | | | | | | * | 0.40 | * | 0.44 | | | | | | | | | | |
| chroma6’, μ | | | | | | | | | * | 0.51 | | | | | | | | | | |
| chroma7’, μ | | | | | | | | | | | | | | | | | | | | |
| chroma8’, μ | | | | | | | | | | | | | | | | | | | | |
| chroma9’, μ | | | | | | | | | | | | | | | | | | | | |
| chroma10’, μ | | | | | | | | | | | | | | | | | | | | |
| chroma11’, μ | | | | | | | | | | | | | | | | | | | | |
| chroma11’, μ | | | | | | | | | | | | | | | | | | | | |
| chroma1’, σ | | | | | * | -0.44 | | | | | | | | | | | | | | |
| chroma2’, σ | | | | | | | | | * | 0.34 | | | | | * | 0.63 | | | | |
| chroma3’, σ | | | | | | | | | | | | | | | * | 0.83 | | | * | 1.99 |
| chroma4’, σ | * | 0.54 | | | | | | | | | | | | | | | | | | |
| chroma5’, σ | | | | | | | | | | | | | | | | | | | | |
| chroma6’, σ | | | | | | | | | * | 0.37 | | | | | | | | | | |
| chroma7’, σ | | | * | -0.36 | | | * | -0.45 | | | ** | 0.69 | * | -0.57 | * | 1.02 | | | | |
| chroma8’, σ | | | * | -0.52 | ** | -0.63 | | | * | 0.38 | | | | | | | | | | |
| chroma9’, σ | | | | | | | | | | | | | | | | | | | | |
| chroma10’, σ | | | | | | | | | | | | | | | | | | | | |
| chroma11’, σ | | | | | | | | | | | | | | | | | | | | |
| chroma12’, σ | | | | | | | | | | | | | | | | | | | | |
| μ chroma’ σ | | | | | | | | | * | 0.32 | | | | | | | | | | |
| slope', m | | | | | | | | | | | | | | | | | | | | |
| slope', s | | | | | | | | | | | ** | -0.35 | | | | | | | | |
| flux', m | | | | | | | | | | | | | | | | | | | | |
| flux', s | | | | | | | | | * | -0.04 | * | -0.30 | | | | | | | | |
| entropy', m | | | | | | | * | -0.42 | * | -0.30 | | | | | | | | | | |
| entropy', s | * | 0.54 | * | 0.42 | | | | | ** | -0.54 | * | -0.61 | | | | | | | | |
| centroid', m | | | | | | | * | -0.35 | * | -0.20 | | | | | | | | | | |
| centroid', s | * | 0.79 | ** | 0.60 | | | | | | | * | -0.47 | | | | | | | | |
| spread', m | | | | | | | * | -0.43 | * | -0.28 | | | | | | | | | | |
| spread', s | ** | 0.76 | * | 0.48 | | | | | | | | | | | | | | | | |
| skewness', m | | | | | | | ** | 0.47 | * | 0.30 | | | | | | | | | | |
| skewness', s | * | 0.51 | | | | | | | * | -0.23 | | | ** | -0.48 | | | | | | |
| kurtosis', m | | | | | | | ** | 0.53 | * | 0.35 | | | | | | | | | | |
| kurtosis', s | | | | | | | | | | | | | ** | -0.59 | | | | | | |
| flatness', m | | | | | | | | | | | | | | | | | | | | |
| flatness', s | | | * | 0.43 | * | 0.44 | | | | | | | | | * | -0.66 | | | | |
| rolloff', m | | | | | | | * | -0.42 | * | -0.26 | | | | | | | | | | |
| rolloff', s | ** | 0.75 | ** | 0.59 | | | | | * | -0.28 | * | -0.52 | | | | | | | | |
| rms', m | | | | | | | | | | | | | | | | | | | | |
| rms', s | | | | | | | | | * | -0.35 | * | -0.40 | | | | | | | | |
| crest_factor', m | | | | | | | | | | | | | | | | | | | | |
| crest_factor', s | | | | | | | | | | | ** | -0.45 | | | | | | | | |
| f0_contour', m | | | | | | | | | | | | | | | | | | | | |
| f0_contour', s | | | | | | | | | | | | | | | | | | | | |
| kurtosis', m | | | | | | | | | | | | | | | | | | | | |
| kurtosis', s | | | | | * | 0.34 | | | | | * | -0.44 | | | | | | | | |
| Shannon E', m | | | | | | | | | | | | | | | | | | | | |
| Shannon E', s | | | | | | | | | * | -0.21 | * | -0.33 | | | | | | | * | -1.00 |

| Feature | Cross-Sectional | | | | | | Longitudinal | | | | | | | | | | | | | |
|---|---|---|---|---|---|---|---|---|---|---|---|---|---|---|---|---|---|---|---|---|
| | vs Ataxic (N=395) | | vs NOSA (N=240) | | vs pre-ataxic (N=199) | | Progression (N=97) | | Δ in Controls (N=43) | | Δ in Ataxia (N=54) | | Progression (N=56) | | Δ in NOSA (N=13) | | Progression (N=48) | | Δ in preataxic (N=5) | |
| | p | d | p | d | p | d | p | d | p | MSDR | p | MSDR | p | d | p | MSDR | p | d | p | MSDR |
| MFCC1’’, μ | | | | | | | * | -0.34 | | | | | | | | | | | | |
| MFCC2’’, μ | | | | | | | | | | | | | | | | | | | | |
| MFCC3’’, μ | | | * | 0.32 | | | | | | | | | | | | | | | | |
| MFCC4’’, μ | | | | | | | | | | | | | | | | | | | | |
| MFCC5’’, μ | | | | | | | | | | | | | | | | | | | | |
| MFCC6’’, μ | | | | | | | | | | | | | | | | | | | | |
| MFCC7’’, μ | | | | | | | | | | | | | | | | | | | | |
| MFCC8’’, μ | | | | | | | | | | | | | * | -0.68 | * | 0.57 | | | | |
| MFCC9’’, μ | | | | | | | | | | | | | | | | | | | | |
| MFCC10’’, μ | | | | | | | | | | | | | | | | | | | | |
| MFCC11’’, μ | | | | | | | | | | | | | | | | | | | | |
| MFCC12’’, μ | | | | | | | | | | | | | | | | | | | | |
| MFCC13’’, μ | | | | | | | | | | | | | | | | | | | | |
| MFCC1’’, σ | | | * | 0.37 | | | | | | | * | -0.48 | | | | | | | | |
| MFCC2’’, σ | * | 0.53 | | | | | | | | | * | -0.47 | | | | | | | | |
| MFCC3’’, σ | * | 0.55 | ** | 0.51 | * | 0.43 | | | * | -0.33 | ** | -0.79 | | | | | | | | |
| MFCC4’’, σ | | | | | | | | | | | | | | | | | | | | |
| MFCC5’’, σ | | | | | | | | | * | 0.35 | | | | | ** | 0.87 | | | * | 1.44 |
| MFCC6’’, σ | | | | | | | ** | 0.24 | | | | | * | 0.44 | | | ** | 0.65 | | |
| MFCC7’’, σ | * | 0.67 | | | | | | | | | | | | | | | | | | |
| MFCC8’’, σ | * | 0.57 | * | 0.32 | * | 0.42 | | | | | | | | | | | | | | |
| MFCC9’’, σ | * | 0.46 | | | | | | | | | | | | | | | | | | |
| MFCC10’’, σ | * | 0.50 | | | | | | | | | | | | | | | | | | |
| MFCC11’’, σ | * | 0.46 | | | | | | | | | | | | | | | | | | |
| MFCC12’’, σ | | | | | * | -0.45 | | | | | | | | | | | | | | |
| MFCC13’’, σ | | | | | | | | | | | | | | | | | | | | |
| μMFCC’’σ | * | 0.57 | * | 0.40 | | | | | | | ** | -0.59 | | | | | | | | |
| chroma1’’, μ | | | | | | | | | | | | | | | | | | | | |
| chroma2’’, μ | | | | | | | | | | | | | | | | | | | | |
| chroma3’’, μ | | | | | | | | | | | | | | | | | | | | |
| chroma4’’, μ | | | | | | | * | -0.33 | | | | | | | | | | | | |
| chroma5’’, μ | | | | | | | ** | -0.39 | | | | | * | -0.49 | | | | | | |
| chroma6’’, μ | | | | | | | | | | | | | | | | | | | | |
| chroma7’’, μ | | | | | | | | | | | | | | | | | | | | |
| chroma8’’, μ | | | | | | | | | | | | | | | | | | | | |
| chroma9’’, μ | | | | | | | | | | | | | | | | | | | | |
| chroma10’’, μ | | | | | | | | | | | | | | | | | | | | |
| chroma11’’, μ | | | | | | | | | | | | | | | | | | | | |
| chroma11’’, μ | | | | | | | | | | | | | | | | | | | | |
| chroma1’’, σ | | | | | * | -0.48 | | | * | 0.40 | | | | | | | | | | |
| chroma2’’, σ | | | | | | | | | | | | | | | | | | | | |
| chroma3’’, σ | | | | | | | | | | | | | | | | | | | * | 1.15 |
| chroma4’’, σ | * | 0.50 | | | | | | | | | | | | | | | | | | |
| chroma5’’, σ | | | | | | | | | | | | | | | | | | | | |
| chroma6’’, σ | | | * | -0.37 | | | | | ** | 0.32 | * | 0.54 | | | * | 0.79 | | | | |
| chroma7’’, σ | | | * | -0.53 | | | ** | -0.50 | | | ** | 0.73 | | | * | 0.85 | | | | |
| chroma8’’, σ | | | * | -0.61 | ** | -0.76 | | | * | 0.41 | * | 0.57 | | | | | | | | |
| chroma9’’, σ | | | | | | | | | | | | | | | | | | | | |
| chroma10’’, σ | | | | | | | | | | | | | | | | | | | | |
| chroma11’’, σ | | | | | | | | | | | | | | | | | | | | |
| chroma12’’, σ | | | | | | | | | | | | | | | | | | | | |
| μ chroma’’ σ | | | | | | | | | | | | | | | | | | | | |
| slope", m | | | | | | | | | | | | | | | * | -0.69 | | | | |
| slope", s | | | | | | | | | | | * | -0.34 | | | | | | | | |
| flux", m | | | | | | | * | -0.50 | | | | | | | | | | | | |
| flux", s | | | | | | | | | * | 0.00 | * | -0.29 | | | | | | | | |
| entropy",m | | | | | | | | | | | | | | | | | | | | |
| entropy", s | * | 0.53 | * | 0.45 | | | | | ** | -0.47 | * | -0.51 | | | | | | | | |
| centroid", m | | | | | | | | | | | | | | | | | | | | |
| centroid", s | * | 0.71 | * | 0.61 | | | | | | | | | | | | | | | | |
| spread", m | | | | | | | | | | | | | | | | | | | | |
| spread", s | * | 0.66 | * | 0.46 | | | | | | | | | | | | | | | * | 1.44 |
| skewness", m | | | | | | | | | | | | | | | | | | | | |
| skewness", s | * | 0.47 | | | | | | | | | | | ** | -0.52 | | | ** | -0.48 | * | 1.08 |
| kurtosis", m | | | | | | | | | | | | | | | | | | | | |
| kurtosis", s | | | | | | | | | | | | | ** | -0.64 | * | 0.56 | ** | -0.57 | | |
| flatness", m | | | | | | | | | | | | | | | | | | | | |
| flatness", s | | | * | 0.45 | * | 0.46 | | | | | | | | | * | -0.65 | | | | |
| rolloff", m | | | | | | | | | | | | | | | | | | | | |
| rolloff", s | * | 0.69 | ** | 0.58 | | | | | * | -0.25 | * | -0.50 | | | | | | | | |
| rms", m | | | | | | | | | | | | | | | | | | | | |
| rms", s | | | | | | | | | ** | -0.37 | * | -0.43 | | | | | | | * | -1.40 |
| crest_factor", m | | | | | | | | | | | | | | | | | | | | |
| crest_factor", s | | | | | | | | | | | ** | -0.43 | | | | | | | | |
| f0_contour", m | | | | | | | | | | | | | | | | | | | | |
| f0_contour", s | | | | | | | | | | | | | | | | | | | | |
| kurtosis", m | | | | | | | | | | | | | | | | | | | | |
| kurtosis", s | | | | | | | | | | | * | -0.41 | | | | | | | * | 1.32 |
| Shannon E", m | | | | | | | | | | | | | | | | | | | | |
| Shannon E", s | * | -0.32 | | | | | | | * | -0.21 | * | -0.34 | | | | | | | * | -0.85 |

*Supplementary Table 9 Correlation between speech-derived features and clinical measures*

| | BARS | | | SARA | | MICARS | | | PROM ataxia | | | | DIS | CPIB |
|---|---|---|---|---|---|---|---|---|---|---|---|---|---|---|
| Feature | Speech | Oculomotor | Total | Speech | Total | Fluency | Clarity | Alternating | Speech | Motor | Comm. | Total | Total | Total |
| MFCC1, μ | 0.28 * | | | 0.28 * | 0.30 * | 0.18 | 0.32 * | 0.27 * | 0.29 * | 0.23 * | 0.26 * | 0.28 * | | |
| MFCC2, μ | | | | | | | | | | | | | | |
| MFCC3, μ | | | | | | | | | | | | | | |
| MFCC4, μ | | | | | | | | | | | | | | |
| MFCC5, μ | | | | | | | | | | | | | | |
| MFCC6, μ | | | | | | | | | | | | | | |
| MFCC7, μ | | | | | | | | | | | | | | |
| MFCC8, μ | | | | | | | | | | | | | | |
| MFCC9, μ | | | | | | | | | | | | | | |
| MFCC10, μ | | | | | | | | | | | | | | |
| MFCC11, μ | | | | | | | | | | | | | | |
| MFCC12, μ | | | | | | | | | | | | | | |
| MFCC13, μ | | | | | | | | | | | | | | |
| MFCC1, σ | 0.29 * | 0.19 * | | | | 0.23 * | 0.29 * | | | | | | | |
| MFCC2, σ | | | | | | | | | | | | | | |
| MFCC3, σ | | 0.16 * | | | | | | | | | | | | |
| MFCC4, σ | | | | | | | | | | | | | | |
| MFCC5, σ | | 0.15 * | | | | | | | | | | | | |
| MFCC6, σ | | | | | | | | | | | | | | |
| MFCC7, σ | | | | | | | | | | | | | | |
| MFCC8, σ | | 0.16 * | | | | | | | | | | | | |
| MFCC9, σ | | | | | | | | | | | | | | |
| MFCC10, σ | | | | | | | | | | | | | | -0.23 * |
| MFCC11, σ | | 0.19 * | | | 0.29 * | | | | | 0.25 * | | | | |
| MFCC12, σ | | | | | | 0.24 * | 0.30 * | 0.26 * | | 0.24 * | | | -0.26 * | |
| MFCC13, σ | | | | | | | | | | | | | | |
| Mean MFCC σ | | 0.17 * | | | | | | | | | | | | |
| MFCC1’, μ | | | | | | | | | | | | | | |
| MFCC2’, μ | | | | | | | | | | | | | | |
| MFCC3’, μ | | | | | | | | | | | | | | |
| MFCC4’, μ | | | | | | | | | | | | | | |
| MFCC5’, μ | | | | | | | | | | | | | | |
| MFCC6’, μ | | | | | | | | | | | | | | |
| MFCC7’, μ | | | | | | | | | | | | | | |
| MFCC8’, μ | | | | | | | | | | | | | | |
| MFCC9’, μ | | | | | | | | | | | | | | |
| MFCC10’, μ | | | | | | | | | | | | | | |
| MFCC11’, μ | | | | | | | | | | | | | | |
| MFCC12’, μ | | | | | | | | | | | | | | |
| MFCC13’, μ | | | | | | | | | | | | | | |
| MFCC1’, σ | | | | | | | | | | | | | | |
| MFCC2’, σ | -0.31 * | | -0.30 * | -0.32 * | | -0.32 * | -0.27 | -0.27 * | -0.25 * | -0.23 * | -0.34 * | -0.25 * | 0.25 * | 0.20 * |
| MFCC3’, σ | | | | | | | | | | | | | | |
| MFCC4’, σ | | | | | | | | | -0.28 * | -0.23 * | -0.37 ** | -0.27 * | 0.37 ** | 0.31 * |
| MFCC5’, σ | | | | | | | | | | -0.22 * | -0.31 * | -0.25 * | 0.30 * | 0.26 * |
| MFCC6’, σ | | | | | | | | | | | | | | 0.29 * |
| MFCC7’, σ | -0.37 * | | -0.35 * | -0.38 * | -0.32 * | -0.29 * | | -0.28 * | -0.25 * | -0.25 * | -0.31 * | -0.30 * | 0.27 * | 0.23 * |
| MFCC8’, σ | -0.32 * | | | -0.32 * | | -0.26 * | -0.29 * | | | | | | 0.25 * | 0.26 * |
| MFCC9’, σ | -0.32 * | -0.20 * | -0.34 * | -0.33 * | -0.29 * | -0.27 * | -0.31 * | | -0.26 * | -0.30 ** | -0.30 * | -0.31 * | | 0.28 * |
| MFCC10’, σ | -0.46 ** | -0.16 * | -0.37 * | -0.46 ** | -0.35 * | -0.34 * | -0.40 ** | -0.42 ** | -0.25 * | -0.24 * | -0.27 * | -0.27 * | 0.33 * | 0.20 * |
| MFCC11’, σ | -0.33 * | | | -0.32 * | | -0.26 * | | -0.30 * | -0.31 * | | -0.32 * | | 0.37 * | 0.30 * |
| MFCC12’, σ | -0.37 * | -0.24 ** | -0.35 * | -0.38 * | -0.34 * | -0.31 * | -0.33 * | -0.33 * | -0.37 ** | -0.24 * | -0.34 * | -0.28 * | 0.34 * | 0.32 ** |
| MFCC13’, σ | -0.36 * | -0.23 ** | -0.32 * | -0.35 * | -0.31 * | -0.25 * | -0.35 * | -0.31 * | -0.26 * | -0.24 * | -0.33 * | -0.29 * | 0.31 * | 0.27 * |
| Mean MFCC’ σ | | | | | | | | | | | -0.28 * | | 0.25 * | 0.23 * |
| MFCC1’’, μ | | | | | | | | | | | | | | |
| MFCC2’’, μ | | | | | | | | | | | | | | |
| MFCC3’’, μ | | | | | | | | | | | | | | |
| MFCC4’’, μ | | | | | | | | | | | -0.22 * | | | 0.21 * |
| MFCC5’’, μ | | | | | | | | | | | | | | |
| MFCC6’’, μ | | | | | | | | | | | | | | |
| MFCC7’’, μ | | | | | | | | | | | | | | |
| MFCC8’’, μ | | | | | | | | | | | | | | |
| MFCC9’’, μ | | | | | | | | | | | | | | |
| MFCC10’’, μ | | | | | | | | | | | | | | |
| MFCC11’’, μ | | | | | | | | | | | | | | |
| MFCC12’’, μ | | | | | | | | | | | | | | |
| MFCC13’’, μ | | | | | | | | | | | | | | |
| MFCC1’’, σ | | | | | | | | | | | | | | |
| MFCC2’’, σ | | | | | | -0.27 * | | | | | -0.27 * | | | |
| MFCC3’’, σ | | | | | | | | | | | | | | |
| MFCC4’’, σ | | | | | | | | | -0.25 * | | -0.32 * | | 0.34 * | 0.26 * |
| MFCC5’’, σ | | | | | | | | | | -0.22 * | -0.32 * | -0.26 * | 0.30 * | 0.25 * |
| MFCC6’’, σ | | | | | | | | | | | | | | 0.25 * |
| MFCC7’’, σ | -0.40 * | -0.17 * | -0.38 ** | -0.40 * | -0.36 * | -0.31 * | -0.33 * | -0.30 * | -0.30 * | -0.30 * | -0.35 * | -0.34 ** | 0.31 * | 0.23 * |
| MFCC8’’, σ | -0.34 * | | -0.30 * | -0.36 * | | -0.29 * | -0.34 * | -0.28 * | -0.26 * | | -0.22 * | | 0.32 * | 0.31 * |
| MFCC9’’, σ | -0.32 * | -0.22 ** | -0.36 * | -0.33 * | -0.32 * | -0.28 * | -0.35 * | -0.25 | -0.27 * | -0.29 * | -0.24 * | -0.29 * | 0.25 * | 0.25 * |
| MFCC10’’, σ | -0.45 ** | -0.22 * | -0.38 ** | -0.45 ** | -0.38 * | -0.30 * | -0.41 ** | -0.43 ** | -0.31 * | -0.24 * | -0.24 * | -0.27 * | 0.34 * | 0.18 * |
| MFCC11’’, σ | -0.32 * | -0.15 * | -0.27 * | -0.31 * | | -0.22 * | -0.28 | -0.28 * | -0.32 * | | -0.28 * | -0.23 * | 0.34 * | 0.25 * |

| | | | | | | | | | | | | | | |
|---|---|---|---|---|---|---|---|---|---|---|---|---|---|---|
| MFCC12'', σ | -0.37 * | -0.25 ** | -0.36 * | -0.37 * | -0.36 * | -0.30 * | -0.36 * | -0.34 * | -0.40 ** | -0.27 * | -0.32 * | -0.30 * | 0.38 ** | 0.32 ** |
| MFCC13'', σ | -0.33 * | -0.22 * | -0.30 * | -0.33 * | -0.30 * | -0.21 | -0.34 * | -0.29 * | -0.27 * | -0.21 * | -0.26 * | -0.25 * | 0.30 * | 0.23 * |
| Mean MFCC'' σ | | | | | | | | | | | -0.29 * | | 0.26 * | 0.22 * |

| | BARS | | | SARA | | MICARS | | | PROM ataxia | | | | DIS | CPIB |
|---|---|---|---|---|---|---|---|---|---|---|---|---|---|---|
| Feature | Speech | Oculomotor | Total | Speech | Total | Fluency | Clarity | Alternating | Speech | Motor | Comm. | Total | Total | Total |
| chroma1, μ | | | | | | | | | | | | | | |
| chroma2, μ | | | | | | | | | | | | | | |
| chroma3, μ | | | | | | | | | | | | | | |
| chroma4, μ | | | | | | -0.23 * | | | | | | | 0.24 * | |
| chroma5, μ | -0.29 * | -0.16 * | -0.31 * | -0.28 * | -0.33 * | -0.24 * | | | -0.25 * | -0.23 * | | | | |
| chroma6, μ | | -0.16 * | | | | | | | -0.27 * | | -0.23 * | | | 0.19 * |
| chroma7, μ | | | | | | | | | -0.26 * | | | | | |
| chroma8, μ | | | | | | | | | | | | | | |
| chroma9, μ | | | | | | | | | | | | | | |
| chroma10, μ | | | | | | | | | | | | | | |
| chroma11, μ | | | | | | | | | | | | | | |
| chroma12, μ | | | | | | | | | | | | | | |
| chroma1, σ | | | | | | | | | | | 0.22 * | | -0.28 * | -0.19 * |
| chroma2, σ | | | | | | | | | 0.32 * | 0.25 * | 0.24 * | 0.25 * | -0.26 * | -0.25 * |
| chroma3, σ | | | | | | | | | | 0.21 * | | | | |
| chroma4, σ | | | | | | | | | | | | | | |
| chroma5, σ | | | | | | | | | | | | | | |
| chroma6, σ | | | | | | | | | | | | | | |
| chroma7, σ | | | | | | | | | | | | | | |
| chroma8, σ | | | | | | | | | | | | | | |
| chroma9, σ | | | | | | | | | | | | | | |
| chroma10, σ | | | | | | | | | | | | | -0.28 * | -0.23 * |
| chroma11, σ | | | | | | | | | | | | | | |
| chroma12, σ | | | | | | | | | | | | | | |
| μ chroma σ | | | | | 0.29 * | | | | 0.32 * | | | | -0.29 * | |
| chroma1', μ | | | | | | | | | | | | | | |
| chroma2', μ | | | | | | | | | | | | | | |
| chroma3', μ | | | | | | | | | | | | | | |
| chroma4', μ | | | | | | | | | | | | | | |
| chroma5', μ | | | | | | | | | | | | | | |
| chroma6', μ | | | | | | | | | | | | | | |
| chroma7', μ | | | | | | | | | | | | | | |
| chroma8', μ | | | | | | | | | | | | | | |
| chroma9', μ | | | | | | | | | | | | | | |
| chroma10', μ | | | | | | | | | | | | | | |
| chroma11', μ | | | | | | | | | | | | | | |
| chroma12', μ | | | | | | | | | | | | | | |
| chroma1', σ | | -0.18 * | | | | | | | | | | | | |
| chroma2', σ | | | | | | | | | | | | | | |
| chroma3', σ | | -0.16 * | | | | | | | | | | | | |
| chroma4', σ | -0.35 * | -0.17 * | -0.32 * | -0.37 * | -0.31 * | -0.31 * | -0.40 ** | -0.35 * | | | | * | 0.28 * | |
| chroma5', σ | -0.34 * | -0.20 * | -0.33 * | -0.35 * | -0.32 * | -0.34 * | -0.37 * | -0.34 * | | | | * | 0.25 * | 0.19 * |
| chroma6', σ | -0.33 * | -0.22 * | -0.31 * | -0.33 * | -0.31 * | -0.35 ** | -0.36 * | -0.33 * | | | | | | |
| chroma7', σ | -0.31 * | | -0.28 * | -0.32 * | -0.31 * | -0.30 * | -0.34 * | -0.31 * | | | | | | |
| chroma8', σ | -0.37 * | -0.22 * | -0.36 * | -0.37 * | -0.38 * | -0.36 ** | -0.39 * | -0.34 * | | -0.26 * | | -0.25 * | 0.24 * | |
| chroma9', σ | | -0.24 ** | | | | -0.24 * | | | | | | | | |
| chroma10', σ | | -0.21 * | | | | | | | | | | | | |
| chroma11', σ | -0.32 * | -0.21 * | -0.33 * | -0.31 * | -0.33 * | -0.31 * | -0.35 * | -0.34 * | | -0.21 * | | -0.24 * | 0.30 * | 0.19 * |
| chroma12', σ | | -0.19 * | -0.28 * | | | -0.24 * | -0.29 * | -0.32 * | | | | | | |
| μ chroma' σ | -0.34 * | -0.22 * | -0.33 * | -0.35 * | -0.32 * | -0.31 * | -0.37 * | -0.33 * | | | | | | |
| chroma1'', μ | | | | | | | | | | | | | | |
| chroma2'', μ | | | | | | | | | | | | | | |
| chroma3'', μ | | | | | | | | | | | | | | |
| chroma4'', μ | | | | | | | | | | | | | | |
| chroma5'', μ | | | | | | | | | | | | | | |
| chroma6'', μ | | | | | | | | | | | | | | |
| chroma7'', μ | | | | | | | | | | | | | | |
| chroma8'', μ | | | | | | | | | | | | | | |
| chroma9'', μ | | | | | | | | | | | | | | |
| chroma10'', μ | | | | | | | | | | | | | | |
| chroma11'', μ | | | | | | | | | | | | | | |
| chroma12'', μ | | | | | | | | | | | | | | |
| chroma1'', σ | | -0.19 * | | | | | | | | | | | | |
| chroma2'', σ | | | | | | | | | | | | | | |
| chroma3'', σ | | | | | | | | | | | | | | |
| chroma4'', σ | -0.28 * | -0.17 * | -0.28 * | -0.31 * | -0.27 | -0.22 * | -0.34 * | -0.29 * | | | | | 0.25 * | |
| chroma5'', σ | -0.32 * | -0.21 * | -0.33 * | -0.34 * | -0.31 * | -0.28 * | -0.36 * | -0.32 * | | -0.21 * | | -0.24 * | 0.26 * | |
| chroma6'', σ | -0.33 * | -0.21 * | -0.32 * | -0.34 * | -0.33 * | -0.33 * | -0.38 * | -0.33 * | | -0.23 * | | -0.23 * | | |
| chroma7'', σ | -0.28 * | | | -0.30 * | -0.29 * | -0.28 * | -0.32 * | -0.28 * | | | | | | |
| chroma8'', σ | -0.33 * | -0.21 * | -0.33 * | -0.34 * | -0.36 * | -0.33 * | -0.36 * | -0.30 * | | -0.25 * | | -0.23 * | | |
| chroma9'', σ | | -0.22 ** | | | | | -0.29 * | | | | | | | |
| chroma10'', σ | | -0.20 * | | | | | | | | | | | | |
| chroma11'', σ | -0.28 * | -0.21 * | -0.28 * | -0.28 | -0.29 * | -0.25 * | -0.32 * | -0.29 * | | | | | 0.24 * | |
| chroma12'', σ | | -0.19 * | | | | | | -0.29 * | | | | | | |
| μ chroma'' σ | -0.31 * | -0.21 * | -0.32 * | -0.33 * | -0.32 * | -0.28 * | -0.36 * | -0.31 * | | | | | | |

| Feature | BARS Speech | BARS Oculomotor | BARS Total | SARA Speech | SARA Total | MICARS Fluency | MICARS Clarity | MICARS Alternating | PROM ataxia Speech | PROM ataxia Motor | PROM ataxia Comm. | PROM ataxia Total | DIS Total | CPIB Total |
|---|---|---|---|---|---|---|---|---|---|---|---|---|---|---|
| slope, μ | | -0.16 * | -0.29 * | -0.28 * | -0.31 * | | -0.30 * | -0.29 * | -0.30 * | | | -0.25 * | | |
| slope, σ | 0.32 * | 0.16 * | 0.34 * | 0.32 * | 0.37 * | | 0.34 * | 0.32 * | 0.35 ** | 0.26 * | 0.23 * | 0.28 * | -0.25 * | -0.19 * |
| slope', μ | | | | | | | | | | | | | | |
| slope', σ | | | | | 0.29 * | | | | 0.26 * | 0.18 | | | | |
| slope'', μ | | | | | | | | | | | | | | |
| slope'', σ | | | 0.27 * | | 0.31 * | | 0.29 * | 0.26 * | 0.29 * | | | 0.23 * | | |
| flux, μ | | | | | | | | | 0.25 * | | | | | |
| flux, σ | 0.29 * | | 0.31 * | 0.29 * | 0.34 * | | 0.32 * | 0.28 * | 0.33 * | 0.23 * | | 0.23 * | | |
| flux', μ | | | | | | | | | | | | | | |
| flux', σ | | | | | 0.30 * | | | | 0.27 * | | | | | |
| flux'', μ | | | | | | | | | | 0.23 * | | 0.26 * | | |
| flux'', σ | | | | | | | | | 0.26 * | | | | | |
| entropy, μ | | | | | | | | | | | | | | |
| entropy, σ | | | | | | | | | | | | | | |
| entropy', μ | | | | | | | | | | | | | | |
| entropy', σ | | | | | | | | | | | -0.35 * | | | 0.22 * |
| entropy'',μ | | | | | | | | | | | | | | |
| entropy'', σ | | | | | | | | | | | -0.34 * | | | 0.20 * |
| centroid,μ | | | | | | | | | | | | | | |
| centroid, σ | | | | | | | | | | | | | | |
| centroid', μ | | | | | | | | | | | | | | |
| centroid', σ | | | | | | -0.25 * | | | | | -0.27 * | | | |
| centroid'', μ | | | | | | | | | | | | | | |
| centroid'', σ | | | | | | | | | | | -0.22 * | | | |
| spread, μ | | | | | | | | | | | | | | |
| spread, σ | | 0.15 * | | | | | | | | | | | | |
| spread', μ | | | | | | | | | | | | | | |
| spread', σ | | | -0.31 * | | -0.29 * | -0.29 * | | | -0.34 * | -0.31 ** | -0.39 ** | -0.34 ** | 0.31 * | 0.29 * |
| spread'', μ | | | | | | | | | | | | | | |
| spread'', σ | | | -0.27 * | | | -0.25 * | | | -0.30 * | -0.29 * | -0.35 * | -0.31 ** | 0.27 * | 0.25 * |
| skewness, μ | | | | | | | | | | | | | | |
| skewness, σ | | | | | | | | | | | | | | |
| skewness', μ | | | | | | | | | | | | | | |
| skewness', σ | -0.28 * | | | -0.28 * | -0.24 | -0.26 * | | | -0.29 * | -0.26 * | -0.37 ** | -0.29 * | 0.27 * | 0.25 * |
| skewness'', μ | | | | | | | | | | | | | | -0.18 |
| skewness'', σ | | | | | | -0.26 * | | | -0.29 * | -0.25 * | -0.36 * | -0.29 * | 0.27 * | 0.25 * |
| kurtosis, μ | | | | | | | | | | | | | | |
| kurtosis, σ | | | | | | | | | | | | | | |
| kurtosis', μ | | | | | | | | | | | | | | |
| kurtosis', σ | | | | | | | | | | | -0.28 * | | | |
| kurtosis'', μ | | | | | | | | | | | | | | -0.20 * |
| kurtosis'', σ | | | | | | | | | | | -0.27 * | | | |
| flatness, μ | | | | | | | | | | | | | | |
| flatness, σ | | | | | | | | | | | | | | |
| flatness', μ | | | | | | | | | | | | | | |
| flatness', σ | | | | | | | | | | | | | | |
| flatness'', μ | | | | | | | | | | | | | | |
| flatness'', σ | | | | | | | | | | | | | | |
| rolloff, μ | | | | | | | | | | | | | | |
| rolloff, σ | | | | | | | | | | | | | | |
| rolloff', μ | | | | | | | | | | | | | | |
| rolloff', σ | | | | | | | | | | -0.26 * | -0.35 * | -0.29 * | 0.24 * | 0.20 * |
| rolloff'', μ | | | | | | | | | | | | | | |
| rolloff'', σ | | | | | | | | | | -0.24 * | -0.32 * | -0.26 * | | |

| Feature | BARS Speech | BARS Oculomotor | BARS Total | SARA Speech | SARA Total | MICARS Fluency | MICARS Clarity | MICARS Alternating | PROM ataxia Speech | PROM ataxia Motor | PROM ataxia Comm. | PROM ataxia Total | DIS Total | CPIB Total |
|---|---|---|---|---|---|---|---|---|---|---|---|---|---|---|
| rms, μ | 0.28 * | | 0.30 * | 0.29 * | 0.34 * | | 0.31 * | 0.31 * | 0.28 * | 0.21 * | | 0.24 * | | |
| rms, σ | 0.33 * | | 0.36 * | 0.34 * | 0.40 ** | | 0.36 * | 0.33 * | 0.35 * | 0.27 * | 0.23 * | 0.29 * | -0.27 * | -0.19 * |
| rms', μ | | | | | | | | | | | | | | |
| rms', σ | | | 0.30 * | 0.28 * | 0.34 * | | 0.31 * | 0.28 * | 0.28 * | 0.21 * | | | | |
| rms'', μ | | | | | | | | | | | | | | |
| rms'', σ | 0.29 * | | 0.31 * | 0.30 * | 0.36 * | | 0.32 * | 0.30 * | 0.28 * | 0.22 * | | 0.23 * | | |
| crest_factor, μ | | | | | | | | | | | | | | |
| crest_factor, σ | | | | | | | | | | | | | | |
| crest_factor', μ | | | | | | | | | | | | | | |
| crest_factor', σ | | | | | | | | | | | | | | |
| crest_factor'', μ | | | | | | | | | | | | | | |
| crest_factor'', σ | | | | | | | | | | | | | | |
| f0_contour, μ | | | | | | 0.25 * | | | | | | | | |
| f0_contour, σ | | | | | | | | | | | | | | |
| f0_contour', μ | | | | | | | | | | | | | | |
| f0_contour', σ | | | | | | | | | | | | | | |

| | | | | | | | | | | | | | | | | | | | | | | | | | | | | |
|---|---|---|---|---|---|---|---|---|---|---|---|---|---|---|---|---|---|---|---|---|---|---|---|---|---|---|---|---|
| f0_contour", μ | | | | | | | | | | | | | | | | | | | | | | | | | | | | |
| f0_contour", σ | | | | | | | | | | | | | | | | | | | | | | | | | | | | |
| kurtosis, μ | | | | | | | | | | | | | | | | | | | | | | | | | | | | |
| kurtosis, σ | | | | | | | | | | | | | | | | | | | | | | | | | | | | |
| kurtosis', μ | | | | | | | | | | | | | | | | | | | | | | | | | | | | |
| kurtosis', σ | | | | | | | | | | | | | | | | | | | | | | | | | | | | |
| kurtosis", μ | | | | | | | | | | | | | | | | | | | | | | | | | | | | |
| kurtosis", σ | | | | | | | | | | | | | | | | | | | | | | | | | | | | |
| Shannon E, μ | 0.30 | * | | | 0.32 | * | 0.31 | * | 0.36 | * | | | 0.32 | * | 0.32 | * | 0.30 | * | 0.23 | * | | | 0.26 | * | | | | |
| Shannon E, σ | 0.32 | * | | | 0.34 | * | 0.34 | * | 0.39 | ** | | | 0.35 | * | 0.32 | * | 0.34 | * | 0.27 | * | 0.23 | * | 0.29 | * | -0.27 | * | -0.19 | * |
| Shannon E, μ | | | | | | | | | | | | | | | | | | | | | | | | | | | | |
| Shannon E', σ | 0.30 | * | | | 0.32 | * | 0.31 | * | 0.36 | * | | | 0.33 | * | 0.30 | * | 0.33 | * | 0.24 | * | | | 0.25 | * | | | | |
| Shannon E", μ | | | | | | | | | | | | | | | | | | | | | | | | | | | | |
| Shannon E", σ | 0.30 | * | | | 0.31 | * | 0.31 | * | 0.37 | * | | | 0.33 | * | 0.30 | * | 0.32 | * | 0.25 | * | 0.22 | * | 0.26 | * | | | | |
| Speaking Rate | -0.67 | ** | -0.23 | ** | -0.67 | ** | -0.63 | ** | -0.68 | ** | -0.61 | ** | -0.61 | ** | -0.58 | ** | -0.47 | ** | -0.47 | ** | -0.43 | ** | -0.44 | ** | 0.53 | ** | 0.43 | ** |
| PD, μ | 0.62 | ** | 0.23 | ** | 0.63 | ** | 0.58 | ** | 0.63 | ** | 0.56 | ** | 0.55 | ** | 0.53 | ** | 0.45 | ** | 0.46 | ** | 0.42 | ** | 0.42 | ** | -0.51 | ** | -0.43 | ** |
| PD, σ | 0.47 | ** | 0.17 | * | 0.49 | ** | 0.43 | ** | 0.46 | ** | 0.42 | ** | 0.41 | ** | 0.36 | ** | 0.42 | ** | 0.38 | ** | 0.38 | ** | 0.34 | ** | -0.43 | ** | -0.40 | ** |
| Percent Pause | 0.45 | ** | 0.19 | * | 0.47 | ** | 0.41 | ** | 0.46 | ** | 0.41 | ** | 0.38 | * | 0.37 | ** | 0.34 | * | 0.36 | ** | 0.32 | * | 0.30 | * | -0.38 | ** | -0.35 | ** |
| Pause Events | | | | | | | | | | | -0.24 | * | | | | | | | | | | | | | 0.26 | * | | |
| Speech Duration | 0.62 | ** | 0.18 | * | 0.62 | ** | 0.58 | ** | 0.62 | ** | 0.55 | ** | 0.57 | ** | 0.55 | ** | 0.44 | ** | 0.40 | ** | 0.42 | ** | 0.42 | ** | -0.49 | ** | -0.40 | ** |
| Total Duration | 0.64 | ** | 0.21 | * | 0.64 | ** | 0.60 | ** | 0.65 | ** | 0.57 | ** | 0.58 | ** | 0.56 | ** | 0.44 | ** | 0.45 | ** | 0.43 | ** | 0.43 | ** | -0.48 | ** | -0.40 | ** |
| Total PD | 0.61 | ** | 0.23 | ** | 0.62 | ** | 0.57 | ** | 0.62 | ** | 0.55 | ** | 0.54 | ** | 0.52 | ** | 0.44 | ** | 0.45 | ** | 0.41 | ** | 0.40 | ** | -0.49 | ** | -0.41 | ** |

*Supplementary Table 10 Correlation between slope of digital measures and slope of clinical measures*

| | BARS | | | SARA | | MICARS | | | PROM ataxia | | | | DIS | CPIB |
|---|---|---|---|---|---|---|---|---|---|---|---|---|---|---|
| Feature | Speech | Oculomotor | Total | Speech | Total | Fluency | Clarity | Alternating | Speech | Motor | Comm. | Total | Total | Total |
| MFCC1, μ | | | | | | | | | | | | | | 0.33 ** |
| MFCC2, μ | | | | | | | -0.29 * | | | | | | -0.36 ** | |
| MFCC3, μ | -0.38 ** | | | -0.33 * | | | | | | | | | | |
| MFCC4, μ | | | | | | | | | | | | | | |
| MFCC5, μ | | | | | | | | | | | | | | |
| MFCC6, μ | | | | | | | | | | | | | 0.30 * | |
| MFCC7, μ | | | | | | | | | | | | | | |
| MFCC8, μ | | | | | | | | | | | | | | |
| MFCC9, μ | | | | | | | | | | | | | | |
| MFCC10, μ | | | | | | | | | | | | 0.34 * | | |
| MFCC11, μ | -0.31 * | | -0.27 * | -0.40 ** | | | -0.35 ** | | | 0.37 ** | | 0.31 * | | |
| MFCC12, μ | | | | | | -0.30 ** | | | | | | | | |
| MFCC13, μ | | | | | | | | | | | | | | |
| MFCC1, σ | | | | | | | | | | | | | | |
| MFCC2, σ | | | | | -0.34 ** | | | | | | | | | |
| MFCC3, σ | | | | | | | | | | | | | | |
| MFCC4, σ | | | | | | | | | | | | | | |
| MFCC5, σ | | | | | | | | | | | | | | |
| MFCC6, σ | | | | | | | | | | | | | | |
| MFCC7, σ | | | | | | | | 0.29 * | | | | | | |
| MFCC8, σ | | | | | | | | | | | | | | |
| MFCC9, σ | | | | | | | | | | | | | | |
| MFCC10, σ | | | | | | | | | | | | | | |
| MFCC11, σ | | | | | | | | | | | | | | |
| MFCC12, σ | | | | | | | | | | | | | | |
| MFCC13, σ | | | | | | | | | | | | | | |
| Mean MFCC σ | | | | | | | | | | | | | | |
| MFCC1’, μ | | | | | | | | | | | | | | |
| MFCC2’, μ | | | | | | | | | 0.33 * | | | | | |
| MFCC3’, μ | | 0.33 ** | | | | | -0.39 ** | | | | | | -0.37 ** | |
| MFCC4’, μ | | | | | | | | | | | | | | |
| MFCC5’, μ | | | | | | | | | | | | | | |
| MFCC6’, μ | | | -0.31 * | | -0.30 * | | | -0.28 * | | | | | | |
| MFCC7’, μ | -0.30 * | | -0.30 * | -0.34 * | | | | -0.29 * | | | | | | |
| MFCC8’, μ | | 0.30 ** | | | | | | | -0.42 ** | | -0.30 * | -0.31 * | | |
| MFCC9’, μ | | | | | | | | | 0.29 * | | | | | |
| MFCC10’, μ | | | | | | | | | | | | | | |
| MFCC11’, μ | | | | | | | | | | | | | | |
| MFCC12’, μ | | | | | | | | | | | | | | |
| MFCC13’, μ | | | | | | | | | | | | | | |
| MFCC1’, σ | | | | | | | | | | | | | | |
| MFCC2’, σ | | | | | -0.40 ** | | | | | | | | | |
| MFCC3’, σ | | | | | | | | | | | -0.32 * | | | |
| MFCC4’, σ | | | | | | | | | | | | | | |
| MFCC5’, σ | | | | | | | | | | | | | | |
| MFCC6’, σ | | | | | | | | | | | | | | |
| MFCC7’, σ | | | | | | | | | | | | | 0.31 ** | |
| MFCC8’, σ | | | | | | | | | | | | | 0.36 ** | |
| MFCC9’, σ | | | | | | | | | | | | | | |
| MFCC10’, σ | | | | | 0.32 ** | | | | | | | | 0.30 ** | |
| MFCC11’, σ | | | | | | | | | | | | | 0.29 * | |
| MFCC12’, σ | | | | | | | | | | | | | | |
| MFCC13’, σ | | | | | 0.32 ** | | | | | | -0.34 ** | | | |
| Mean MFCC’ σ | | | | | | | | | | | | | | |
| MFCC1’’, μ | | | | | | | | | | | -0.33 * | -0.29 * | | |
| MFCC2’’, μ | | | | | | | | | | | | | | |
| MFCC3’’, μ | | | | | | | | | | | 0.29 * | | | |
| MFCC4’’, μ | | | | | | | | 0.29 * | | | | | | |
| MFCC5’’, μ | | | | | | | | | | | | | | |
| MFCC6’’, μ | | | | | | | | | | | | | | |
| MFCC7’’, μ | | | | | | | | | | | | | | |
| MFCC8’’, μ | -0.29 * | | | | | | -0.35 ** | -0.37 ** | | | | | | -0.37 ** |
| MFCC9’’, μ | -0.33 * | | | -0.34 * | | | | | | | | 0.32 * | | |
| MFCC10’’, μ | | | | | | | | | | | | | -0.33 ** | |
| MFCC11’’, μ | | | | | | | | | | | | | | |
| MFCC12’’, μ | | | | | | | | | | | | | | |
| MFCC13’’, μ | | | | | | | | | | | | | | |
| MFCC1’’, σ | | | | | | | | | | | -0.30 * | | | |
| MFCC2’’, σ | | | | | -0.37 ** | | | | | | | | | |
| MFCC3’’, σ | | | | | | | | | | | -0.35 ** | | | |

| | | | | | | | | | | | | |
|---|---|---|---|---|---|---|---|---|---|---|---|---|
| MFCC4'', σ | | | | | | | | | | -0.28 * | | |
| MFCC5'', σ | | | | | | | | | | | | |
| MFCC6'', σ | | | | | | | | | | | | |
| MFCC7'', σ | | | | | | | | | | | | 0.29 * |
| MFCC8'', σ | | | | | | | | | | | | 0.38 ** |
| MFCC9'', σ | | | | | -0.28 ** | | | | | | | |
| MFCC10'', σ | | | | 0.38 ** | | | | | -0.31 * | | | |
| MFCC11'', σ | | | | | | | | | | | | |
| MFCC12'', σ | | | | | | | | | | | 0.28 * | |
| MFCC13'', σ | -0.30 * | | | 0.31 * | | | | | | | | |
| Mean MFCC'' σ | | | | | | | | | | -0.32 * | | |

| | BARS | | | SARA | | MICARS | | | PROM ataxia | | | | DIS | CPIB |
|---|---|---|---|---|---|---|---|---|---|---|---|---|---|---|
| Feature | Speech | Oculomotor | Total | Speech | Total | Fluency | Clarity | Alternating | Speech | Motor | Comm. | Total | Total | Total |
| chroma1, μ | | 0.35 ** | | | | | | | | | -0.32 * | | | |
| chroma2, μ | | | | | | | | -0.33 * | | 0.28 * | | | | |
| chroma3, μ | | | | | | | | -0.34 ** | | | | | | |
| chroma4, μ | | | | | | | | | 0.34 * | | 0.36 ** | | | |
| chroma5, μ | | | | | | | | | | | | | | |
| chroma6, μ | | | | | | | | | | | | | | |
| chroma7, μ | | | | | | | | | | | | | | |
| chroma8, μ | | | | | | | | | -0.31 * | | -0.30 * | | | |
| chroma9, μ | | | | | -0.28 * | | | | -0.42 ** | | -0.33 * | | | |
| chroma10, μ | | | | | -0.28 * | | | | -0.38 * | | | | | |
| chroma11, μ | 0.36 ** | | | | | | | | | | | | | |
| chroma12, μ | | 0.36 ** | 0.31 * | | | | | | -0.28 * | | -0.40 ** | | | |
| chroma1, σ | | | | | | | | | | | | | | |
| chroma2, σ | | | | | | | | | | 0.31 * | | 0.38 * | | |
| chroma3, σ | | | | | | | | | | | | 0.39 * | | |
| chroma4, σ | | | | | | | | | | | | | | |
| chroma5, σ | | | | | | | | | | | | | | |
| chroma6, σ | | | | | | | | | | | | | | |
| chroma7, σ | | | -0.29 * | | | | | | | 0.39 ** | | 0.42 * | | |
| chroma8, σ | -0.29 * | | -0.28 * | | | | | | | 0.33 ** | | 0.45 ** | | |
| chroma9, σ | -0.30 * | | -0.41 ** | -0.49 ** | | | | | | | | 0.35 * | | |
| chroma10, σ | | | | | | | | | | | | | | |
| chroma11, σ | | | | | | | | | | | | | | |
| chroma12, σ | | | | | | | | | | 0.32 * | | | | |
| μ chroma σ | | | -0.34 * | | | | | | | 0.32 * | | 0.46 ** | | |
| chroma1', μ | | | | | | | | | | | | | | |
| chroma2', μ | | | | | | | | -0.31 * | | | | | | |
| chroma3', μ | | | | | | | | | | | | | | |
| chroma4', μ | -0.30 * | | -0.32 * | | | | | -0.32 * | | | | | | |
| chroma5', μ | | | | | | | | | | | | | | |
| chroma6', μ | | | | | | | | | | | | | | |
| chroma7', μ | | | | | | | | | | | | | | |
| chroma8', μ | | | | | | | | | | | | | | |
| chroma9', μ | | | | | | | | | | | | | | |
| chroma10', μ | | | | | | | | | | | | | | |
| chroma11', μ | | | | | | | | | | | | | | |
| chroma12', μ | | | | | | | | | | | | | | -0.31 ** |
| chroma1', σ | | 0.39 ** | | | | | | | | | | | | |
| chroma2', σ | | | | | | | | | | 0.28 * | | 0.29 * | | |
| chroma3', σ | -0.28 * | | -0.30 * | -0.38 * | | | | -0.35 ** | | 0.29 * | | 0.34 * | | |
| chroma4', σ | -0.35 * | | -0.41 ** | -0.39 ** | | | | -0.42 ** | | | | 0.31 * | | |
| chroma5', σ | -0.29 * | | -0.39 ** | -0.27 * | | | | | | | | 0.36 * | | |
| chroma6', σ | | | | | | | | | | | | | | |
| chroma7', σ | | | | | | | | | | | | | | 0.34 ** |
| chroma8', σ | | | | | | | | | | | | | | |
| chroma9', σ | | | | | | | | | | | | 0.31 * | | |
| chroma10', σ | | | | -0.34 * | | | | | | | | 0.29 * | | |
| chroma11', σ | | | | | | | | | | | | | | |
| chroma12', σ | | | | | | | | | | | | | | |
| μ chroma' σ | | | | | | | | | | | | | | |
| chroma1'', μ | -0.30 * | | | | | | | | | | | | | -0.29 * |
| chroma2'', μ | | | | | | | | | | | | | | |
| chroma3'', μ | | | | | | | | | | | | | | |
| chroma4'', μ | | | | | | | | | | | | | | |
| chroma5'', μ | | | | 0.37 * | | | | | -0.32 * | | | -0.33 * | | |
| chroma6'', μ | | 0.29 * | | 0.39 ** | | | | | -0.28 * | | | -0.41 * | | |
| chroma7'', μ | | 0.37 ** | | 0.28 * | | | | | -0.29 * | | | -0.29 * | | |
| chroma8'', μ | | 0.29 ** | | | | | | | | | | | | |
| chroma9'', μ | | | | | | | | | | | | | | |
| chroma10'', μ | | | | | | | | | | | | | | |

| | | | | | | | | | | | | | | |
|---|---|---|---|---|---|---|---|---|---|---|---|---|---|---|
| chroma11’’, μ | | | | | | | | | | | | 0.29 * | | |
| chroma12’’, μ | | | | | | | | | | | | | | -0.31 ** |
| chroma1’’, σ | | 0.35 ** | | | | | | | -0.29 * | | | | | |
| chroma2’’, σ | | | | | | | | | | | | | | |
| chroma3’’, σ | -0.31 * | | -0.30 * | -0.40 ** | | | -0.30 * | -0.37 ** | | 0.29 * | | 0.31 * | | |
| chroma4’’, σ | -0.37 ** | | -0.37 * | -0.38 * | | | | -0.41 ** | | | | | | |
| chroma5’’, σ | -0.38 ** | | -0.38 ** | -0.29 * | | | | | | | | 0.35 * | | |
| chroma6’’, σ | | | | | | | | | | | | | | |
| chroma7’’, σ | | | | | | | | | | | | | | 0.31 ** |
| chroma8’’, σ | | | | | | | | | | | | | | |
| chroma9’’, σ | | | | | | | | | | | | | | |
| chroma10’’, σ | | | | -0.28 * | | | | | | | | | | |
| chroma11’’, σ | | | | | | | | | | | | | | |
| chroma12’’, σ | | | | | | | | | | | -0.28 * | | | |
| μ chroma’’ σ | | | | | | | | | | | | | | |

| | BARS | | | SARA | | MICARS | | | PROM ataxia | | | | DIS | CPIB |
|---|---|---|---|---|---|---|---|---|---|---|---|---|---|---|
| Feature | Speech | Oculomotor | Total | Speech | Total | Fluency | Clarity | Alternating | Speech | Motor | Comm. | Total | Total | Total |
| slope, μ | | | | | | | | | | | | | | |
| slope, σ | | | | | -0.29 * | | | | | | | | | |
| slope', μ | | | | | | | | | | | | | | |
| slope', σ | | | | | | | | | | | | | | |
| slope'', μ | | | -0.34 * | | | | | | | | | | | |
| slope'', σ | | | | | | | | | | | | | | |
| flux, μ | | | | | | | | | | | | | | |
| flux, σ | | | | | | | | | | | | | | |
| flux', μ | | | | | | | | | | | | | | |
| flux', σ | | | | | -0.31 ** | | | | | | | | | |
| flux'', μ | | | | | | | | | | | | | | |
| flux'', σ | | | | | -0.30 * | | | | | | | | | |
| entropy, μ | | | | | | | | | | | | 0.36 * | | |
| entropy, σ | | | | | | | | | | | | | | |
| entropy', μ | | | | | | | 0.31 * | | -0.30 * | | | | | |
| entropy', σ | | | | | | | | | | | | | | |
| entropy'',μ | | | | | | | | | | | | | | |
| entropy'', σ | | | | | | | | | | | | | | |
| centroid,μ | | | | | | | | | | | | 0.36 * | | |
| centroid, σ | | | | | | | | | | | | | | |
| centroid', μ | | | | | | | 0.34 ** | | -0.39 ** | | -0.28 * | | | |
| centroid', σ | | | | | | | | | | | | | | |
| centroid'', μ | | | | | | | 0.33 ** | | | | | | | |
| centroid'', σ | | | | | | | | | | | | | | |
| spread, μ | | | | | 0.27 * | | | | | | | 0.30 * | | |
| spread, σ | | | -0.28 * | | | -0.29 ** | | | 0.32 * | | | | | -0.29 ** |
| spread', μ | | | | | | | | | -0.33 * | | | | | |
| spread', σ | | | | | | | | | | | | | | |
| spread'', μ | | | | | | | | | | | | | | |
| spread'', σ | | | | | | | | | | | | | | |
| skewness, μ | | | | | | | | | | | | -0.28 * | | |
| skewness, σ | | | | | | | | | | | | | | |
| skewness', μ | | | | | | | -0.29 * | | 0.41 ** | | 0.28 * | 0.29 * | | |
| skewness', σ | | | | | | | | | | | -0.30 * | -0.41 * | | |
| skewness'', μ | | | | | | | -0.29 * | | | | | | | |
| skewness'', σ | | | | | | | | | | -0.29 * | -0.32 * | -0.44 ** | | |
| kurtosis, μ | | | | | | | | | | | | -0.29 * | | |
| kurtosis, σ | | | | | | | | | | | | | | |
| kurtosis', μ | | | | | | | | | 0.40 ** | 0.28 * | 0.28 * | 0.34 * | | |
| kurtosis', σ | | | | | | | | | | -0.31 * | -0.29 * | -0.44 ** | | |
| kurtosis'', μ | | | | | | | | | | | | | | |
| kurtosis'', σ | | | | | -0.28 * | | | | | -0.33 ** | -0.31 * | -0.47 ** | | |
| flatness, μ | | | | | | | | | | | | 0.37 * | | |
| flatness, σ | | | | | | | | | | | 0.30 * | 0.31 * | | |
| flatness', μ | | | | | | | 0.32 ** | | -0.33 * | | | | | |
| flatness', σ | | | | | | | | | | 0.30 * | | 0.40 * | | |
| flatness'', μ | | | | | | | | | | | | | | |
| flatness'', σ | | | | | | | | | 0.29 * | 0.32 * | 0.28 * | 0.45 ** | | |
| rolloff, μ | | | | | | | | | | | | 0.33 * | | |
| rolloff, σ | | | -0.29 * | | | | | | | | | | | |
| rolloff', μ | | | | | | | 0.34 ** | | -0.40 ** | | -0.33 * | | | |
| rolloff', σ | | | | | | | | | | | | | | |
| rolloff'', μ | | | | | | | 0.31 * | | | | | | | |
| rolloff'', σ | | | | | | | | | | | | | | |

| Feature | BARS Speech | BARS Oculomotor | BARS Total | SARA Speech | SARA Total | MICARS Fluency | MICARS Clarity | MICARS Alternating | PROM ataxia Speech | PROM ataxia Motor | PROM ataxia Comm. | PROM ataxia Total | DIS Total | CPIB Total |
|---|---|---|---|---|---|---|---|---|---|---|---|---|---|---|
| rms, μ | | | | | | | | | | | | | | |
| rms, σ | | | | | | | | | | | | | | |
| rms', μ | | | | | | | | | | | | | | |
| rms', σ | | | | | | | | | | | | | | |
| rms", μ | | | | | | | | | | | | | | |
| rms", σ | | | | | | | | | | | | | | |
| crest_factor, μ | | | | | | | | | | | | | | |
| crest_factor, σ | | | | | -0.28 * | | | | | | | | -0.29 * | |
| crest_factor', μ | | | | | | | | | | | | | | |
| crest_factor', σ | | | | | -0.29 * | | | | | | | | | |
| crest_factor", μ | | | | | | | | | | | | | | |
| crest_factor", σ | | | | | | | | | | | | | | |
| f0_contour, μ | | | | | -0.36 ** | | | | | | | | | |
| f0_contour, σ | | | | | | | | | | 0.35 ** | | | | |
| f0_contour', μ | 0.37 ** | | | | | | | | | -0.29 * | | -0.38 * | | |
| f0_contour', σ | | | | | | | | | | | -0.34 ** | | | |
| f0_contour", μ | | | | | | | | | | 0.33 ** | | 0.34 * | | |
| f0_contour", σ | | | | | | | | | | | -0.34 * | | | |
| kurtosis, μ | | | | -0.30 * | | | -0.28 * | | | | | | | |
| kurtosis, σ | | | | | | | | | | | | | -0.31 ** | |
| kurtosis', μ | | | | | | | | | | | | | | |
| kurtosis', σ | | | | | | | | | | | | | -0.29 * | |
| kurtosis", μ | | | | | | | | | | | | | | |
| kurtosis", σ | | | | | | | | | | | | | -0.30 * | |
| Shannon E, μ | | | | | | | | | | | | | | |
| Shannon E, σ | | | | | | | | | | | | | | |
| Shannon E, μ | | | | | | | | | | | | | | |
| Shannon E', σ | | | | | | | | | | | | | | |
| Shannon E", μ | | | | | | | 0.29 * | | | | | | | |
| Shannon E", σ | | | | | | | | | | | | | | |

| Feature | BARS Speech | BARS Oculomotor | BARS Total | SARA Speech | SARA Total | MICARS Fluency | MICARS Clarity | MICARS Alternating | PROM ataxia Speech | PROM ataxia Motor | PROM ataxia Comm. | PROM ataxia Total | DIS Total | CPIB Total |
|---|---|---|---|---|---|---|---|---|---|---|---|---|---|---|
| Speaking Rate | | | | | | | | | | | | -0.36 * | | |
| PD, μ | | | | | | | | | | | | 0.38 * | | |
| PD, σ | | | | | | | | | | | | | | |
| Percent Pause | | | | | | | | | | | | | | |
| Pause Events | | | | | | | | | | | | | | |
| Speech Duration | | | | | | | | | 0.29 * | | | | | |
| Total Duration | | | | | | | | | 0.32 * | | 0.29 * | 0.39 * | | |
| Total PD | | | | | | | | | 0.34 * | | 0.30 * | 0.43 * | | |
